# Adverse childhood trauma and reoccurrence of illness impact the gut microbiome, which affects suicidal behaviors and the phenome of major depression: towards enterotypic-phenotypes

**DOI:** 10.1101/2023.01.14.23284564

**Authors:** Michael Maes, Asara Vasupanrajit, Ketsupar Jirakran, Pavit Klomkliew, Prangwalai Chanchaem, Chavit Tunvirachaisakul, Kitiporn Plaimas, Apichat Suratanee, Sunchai Payungporn

## Abstract

The first publication demonstrating that major depressive disorder (MDD) is associated with alterations in the gut microbiota appeared in 2008 (Maes et al., 2008). The purpose of the present study is to delineate a) the microbiome signature of the phenome of depression, including suicidal behaviours and cognitive deficits; the effects of adverse childhood experiences (ACE) and recurrence of illness index (ROI) on the microbiome; and the microbiome signature of lowered high-density lipoprotein cholesterol (HDLc).

We determined isometric log-ratio abundances or prevalence of gut microbiome phyla, genera, and species by analyzing stool samples from 37 healthy Thai controls and 32 MDD patients using 16S rDNA sequencing.

Six microbiome taxa accounted for 36% of the variance in the depression phenome, namely *Hungatella* and *Fusicatenibacter* (positive associations) and *Butyricicoccus, Clostridium, Parabacteroides merdae*, and *Desulfovibrio piger* (inverse association). This profile (labeled enterotype 1) indicates compositional dysbiosis, is strongly predicted by ACE and ROI, and is linked to suicidal behaviours. A second enterotype was developed that predicted a decrease in HDLc and an increase in the atherogenic index of plasma (*Bifidobacterium, P. merdae*, and *Romboutsi*a were positively associated, while *Proteobacteria* and *Clostridium sensu stricto* were negatively associated). Together, enterotypes 1 and 2 explained 40.4% of the variance in the depression phenome, and enterotype 1 in conjunction with HDLc explained 39.9% of the variance in current suicidal behaviours.

In conclusion, the microimmuneoxysome is a potential new drug target for the treatment of severe depression and suicidal behaviours, and possibly for the prevention of future episodes.

## Introduction

2008 marked the first publication demonstrating that major depressive disorder (MDD) is associated with alterations in the gut microbiota (Maes et al., 2008). This study demonstrated that serum levels of IgA and IgM directed against the lipopolysaccharides (LPS) of *Pseudomonas putida, Hafnia alvei, Morganella morganii, Citrobacter koseri, Pseudomonas aeruginosa,* and *Klebsiella pneumoniae* were significantly higher in MDD compared to controls (Maes et al., 2008), indicating that a significant proportion of MDD patients exhibit increased translocation of LPS or Gram-negative enterobacteria via increased gut permeability or leaky gut (Maes et al., 2008). Importantly, this increased translocation of LPS or Gram-negative bacteria was strongly associated with numerous inflammatory, immune activation, oxidative stress, and autoimmunity indicators (Maes et al., 2012). The primary findings of Maes et al. (2008) were corroborated by recent findings that MDD is associated with increased gut permeability (as measured by the lactulose/mannitol test), increased levels of *Morganella* and *Klebsiella*, leaky gut biomarkers, and associations between the latter and inflammatory or anti-inflammatory markers, such as T regulatory (Treg) cells (Calarge et al., 2019; Ohlsson et al., 2019; Alvarez-Mon et al., 2021; 2013 Runners-Up, 2013; Iordache et al., 2022; Simeonova et al., 2018).

As a result, it was proposed that increased LPS translocation may be one of the causes of immune activation and oxidative stress in MDD by activating the Toll-Like Receptor-4 (TLR4) complex and, consequently, nuclear factor-κB (NF-κB) (Lucas and Maes, 2013). There is now evidence that MDD is a disorder characterized by activated immune-inflammatory and nitro-oxidative pathways and that these pathways to a large extent determine the MDD phenome and accompanying suicidal behaviors (Maes et al., 1990; 1997; Maes et al., 2021; 2022a; 2002b; Maes, 2022; Vasupanrajit et al., 2021; 2022). Activated immune and oxidative stress pathways may cause epithelial tight junction abnormalities that increase intestinal permeability and bacterial translocation (Maes et al., 2008; 2012). Consequently, there are reciprocal associations between gut microbiota and increased bacterial translocation due to leaky gut, and systemic immune-oxidative pathways and this interconnected system is best referred to as the “microimmuneoxysome”.

Intestinal dysbiosis, specifically the disbalance in the gut microbiome between pathobionts (pro-inflammatory, causing injuries to epithelial cells and tight junctions) and microbiota that promote salutogenesis (including anti-inflammatory activities, support of gut homeostasis and tight junctions, production of short-chain fatty acids (SCFA) and vitamins), is another potential cause of leaky gut and bacterial translocation (Simeonova et al., 2018; Rudzki and Maes, 2020; Slyepchenko et al., 2017). Gut dysbiosis may also contribute to the co-occurrence of MDD and comorbid metabolic disorders such as type 2 diabetes mellitus (T2DM), obesity, and atherosclerosis (Slyepchenko et al., 2016; Agusti et al., 2018).

Using second-generation sequencing of bacterial 16S RNA genes in conjunction with Linear Discriminant Analysis Effect Size (LEfse) analysis, it was discovered that nearly all studies report changes in gut microbiome phyla, genera, or species (Borkent et al., 2022). Nevertheless, the latter systematic review did not reveal consistent changes in microbiome communalities across studies (Borkent et al., 2022). Possibly, one could deduce from the several studies in the latter systematic review that there are maybe alterations in *Lactobacillus, Streptococcus*, *Eggerthella* and *Faecalibacterium* in patients with mental illnesses.

Several factors may account for the inconsistent nature of these results. First, it is remarkable that the majority of authors failed to discuss the results in terms of compositional dysbiosis, leaving the results without any mechanistic explanation. Second, the microbiome is strongly influenced by diet; consequently, region- or culture-specific alterations in the microbiome may define different microbiome profiles of MDD in different countries or cultures (Singh et al., 2017). Last but not least, the diagnosis of MDD is practically useless for biomarker research because MDD is an incorrect outcome variable that can hardly be used in statistical analysis (Maes, 2022; Maes et al., 2022a; Maes and Stoyanov, 2022). Indeed, MDD is a heterogeneous group that includes severe depression, melancholia phenotypes, mild depression, and possibly even normal human emotional responses such as grief, sadness, and despondency (Maes et al., 2022; Maes and Stoyanov, 2022). Moreover, the DSM/ICD diagnostic criteria for MDD are unreliable, with low inter-psychiatrist reproducibility (Maes and Stoyanov, 2022). Furthermore, MDD is a post-hoc, higher-order construct that is limited in scope because it is a flawed binary construct that does not include the major features of depression, such as recurrence of illness (ROI), lifetime and current suicidal behaviors, and the phenome of depression (Maes and Stoyanov, 2022).

We have recently developed a new clinimetrics method, referred to as “precision nomothetic psychiatry”, which allows us to examine the causal links between causome/protectome factors, ROI, cognitive deficits, and a quantitative score of the phenome of depression (Maes, 2022; Maes et al., 2021; 2022b; Maes and Stoyanov, 2022; Simeonova et al., 2021). Our models demonstrate that adverse childhood experiences (ACE) and increased translocation of Gram-negative bacteria are strongly associated with the phenome of depression (conceptualized as latent vectors extracted from symptom domains, suicidal behaviours, etc.) and that these effects are mediated by ROI, lowered antioxidant defenses, including lowered high-density lipoprotein cholesterol (HDLc), and activated immune and oxidative stress pathways (Moraes et al., 2018; Maes et al., 2019; 2021; 2022b; Maes, 2022). It is intriguing that a pilot study discovered that ACE could influence the microbiome composition during pregnancy, thus contributing to systemic inflammatory responses (Hantsoo et al., 2019). However, there are no data indicating whether ACE and ROI may affect the microbiome or whether compositional dysbiosis may mediate the effects of ACE on the phenome of depression, which includes cognitive deficits and suicidal behaviours.

Hence, the present study was conducted to delineate a) the microbiome signature of the phenome of depression, including suicidal behaviors and cognitive deficits; b) the effects of ACE and ROI on the microbiome; c) the microbiome signature of lowered HDLc, as a major marker of antioxidant defenses and increased atherogenicity in depression.

## Materials and Methods

### Participants

We recruited 37 normal controls and 32 MDD patients from the outpatient clinic of the Department of Psychiatry at King Chulalongkorn Memorial Hospital in Bangkok, Thailand. Participants were of both sexes and between the ages of 19 and 58. The control group was recruited through word of mouth in the same catchment area as the patients, Bangkok, Thailand. Depressed patients were given a diagnosis of MDD based on DSM-5 criteria. Participants (patients and controls) with a DSM-5 axis 1 disorder diagnosis other than MDD were excluded from the study, including those with autism, obsessive-compulsive disorder, post-traumatic stress disorder, substance use disorder (except nicotine dependence), bipolar disorder, psycho-organic disorders, schizophrenia, and schizo-affective disorder. In addition, excluded from the study were healthy control participants with any DSM-5 axis 1 disorder diagnosis (see above) and MDD, and a positive family history of MDD, bipolar disorder, or suicide. Furthermore, participants were excluded for medical illness and conditions including: a) neuroinflammatory and neurodegenerative disorders, such as multiple sclerosis, Alzheimer’s and Parkinson’s disease, epilepsy, and stroke; b) immune and autoimmune disorders, such as cancer, diabetes type 1, psoriasis, systemic lupus erythematosus, COPD, inflammatory bowel disease, irritable bowel syndrome; and c) allergic or inflammatory reactions three months prior to the study. In addition, we excluded: a) pregnant or lactating women; b) patients who were ever treated with immunomodulatory drugs like glucocorticoids or immunosuppressive; c) subjects who were treated with pharmaceutical dosages of antioxidants or omega-3 preparates; and d) patients who had suffered from moderate/critical COVID-19 and who had suffered from mild COVID-19 six months prior to enrollment.

Before participating in this study, all participants provided written informed consent. The research was conducted in accordance with international and Thai ethical standards and privacy laws. The Institutional Review Board of the Chulalongkorn University Faculty of Medicine in Bangkok, Thailand (#528/63) approved the study in accordance with the International Guidelines for the Protection of Human Subjects as required by the Declaration of Helsinki, The Belmont Report, the CIOMS Guideline, and the International Conference on Harmonization in Good Clinical Practice (ICH-GCP).

### Clinical assessments

A well-trained research psychologist experienced in the study of affective disorders conducted semi-structured interviews to collect socio-demographic information, such as gender, age, and level of education. The same research psychologist also collected clinical information, including the number of previous depressive episodes, family medical history, medical history, and psychotropic medications. A senior psychiatrist diagnosed MDD utilizing DSM-5 criteria and the Mini International Neuropsychiatric Interview (M.I.N.I.) (Udomratn and Kittirattanapaiboon et al., 2004). The M.I.N.I. was used to evaluate other axis-1 diagnoses and to exclude patients and controls accordingly. The 17-item Hamilton Depression Rating Scale (HDRS) was used by the research psychologist to assess the severity of depressive symptoms (Hamilton, 1960). The Beck Depression Inventory II was used to assess the severity of self-reported depression (BDI-II, Beck et al., 1996). The latter is a 21-item self-report inventory that was translated into Thai by Nantika Thavichachart et al. to assess the presence and severity of depressive symptoms (Thavichachart et al., 2009).

ACEs were measured using a Thai translation of the Adverse Childhood Experiences Questionnaire (Rungmueanporn et al., 2019). This questionnaire consists of 28 questions regarding childhood traumatic experiences. In the present study, we used 5 ACE domains, including emotional abuse (2 items), physical abuse (2 items), sexual abuse (4 items), emotional neglect (5 items), physical neglect (5 items), and used principal component analysis (PCA) as a feature reduction method to compute scores on sexual abuse, emotional neglect, and physical neglect (see below). In addition, we examined whether it was possible to derive PCs from all abuse and neglect symptoms in order to create PC scores that reflect “abuse” and “neglect.” We utilized the Columbia Suicide Severity Rating Scale (C-SSRS) to assess the severity of lifetime and current suicidal ideation and attempts. The C–SSRS was created by Posner et al. (2011). The test measures the severity and intensity of suicidal ideation, attempts, lethality, and self-harm without suicidal intent. We calculated the PCs extracted from lifetime (LT) and current (current) suicidal ideation (SI) and attempts (SA) and suicidal behaviors (SB, ideation and attempts combined) as explained previously (Maes et al., 2022a). As such, we derived scores of lifetime (LT)_SI, LT_SA, LT_SB, current_SI, current_SA, current_SB, and overall SB (a PC extracted from LT and current SI and SA) (Maes et al., 2022a; Maes, 2022). The research psychologist also examined the Stroop color and word test (SCWT), namely parts 1 (a neutral trial that measures reaction times), part 2 (congruent trial) and part 3 (incongruent trial) (Stroop, 1935). We examined whether the first PC could reflect aberrations in the three Stroop subtests. Tobacco use disorder (TUD) was identified and diagnosed using DSM-5 criteria. Metabolic Syndrome (MetS) was diagnosed using the criteria established by the International Diabetes Federation (Alberti et al., 2006). Weight (in kilos) was divided by the person’s squared height (in meters) to determine their body mass index (BMI).

### Assays

Stool sample collection, DNA extraction, 16S rDNA amplification and 16S rDNA amplicon sequencing based on Oxford Nanopore Technology (ONT) were performed as published previously (Maes et al., 2022c). Approximately 20 mg of stool was collected in sterile test tubes containing 2 mL of DNA/RNA ShieldTM reagent (ZYMO Research, USA) and stored at -20 degrees Celsius until analysis. The DNA was extracted using the ZymoBIOMICS^TM^ DNA Miniprep Kit (ZYMO Research, USA) according to the manufacturer’s instructions. “ The full length of the bacterial 16S rDNA gene (1.5 kb) was amplified by PCR using specific primers: 5′-TTTCTGTTGGTGCTGATATTGCAGRGTTYGATYMTGGCTCAG-3′ and 5′- ACTTGCCTGTCGCTCTATCTTCCGGYTACCTTGTTACGACTT-3′ as described previously (Jitvaropas et al., 2022). The first round of PCR reaction contained 1 µg of DNA template, 0.2 µM of each primer, 0.2 mM of dNTPs, 1X Phusion™ Plus buffer, 0.4 U of Phusion Plus DNA Polymerase (Thermo Scientific, USA) and nuclease-free water in a final volume of 20 µL. The PCR reaction was performed under the following thermal conditions: 98°C for 30 s; 25 cycles of amplification (98°C for 10 s, 60°C for 25 s, 72°C for 45 s) and followed by 72°C for 5 min. After that, the barcodes were attached to the 16S rDNA amplicon by 5 cycles of amplification (98°C for 10 s, 60°C for 25 s, 72°C for 45 s) based on PCR Barcoding Expansion 1–96 (EXP-PBC096) kit (Oxford Nanopore Technologies, UK). The amplicons were purified using QIAquick^®^ PCR Purification Kit (QIAGEN, Germany) according to the manufacturer’s protocol. The concentrations of purified amplicons were measured using a Qubit 4 fluorometer with Qubit dsDNA HS Assay Kit (Thermo Scientific, USA). Then the amplicons with different barcodes were pooled at equal concentrations and purified using 0.5X Agencourt AMPure XP beads (Beckman Coulter, USA). After that, the purified DNA library was end-repaired and adaptor-ligated using Ligation Sequencing Kit (SQK-LSK112) (Oxford Nanopore Technologies, UK). Finally, the library was sequenced by the MinION Mk1C platform with R10.4 flow cell (Oxford Nanopore Technologies, UK).

Guppy basecaller software v6.0.7 (Wick et al., 2019) (Oxford Nanopore Technologies, UK) was used for base-calling with a super-accuracy model to generate pass reads (FASTQ format) with a minimum acceptable quality score (Q > 10). The quality of reads was examined by MinIONQC (Lanfear et al., 2019). Then, FASTQ sequences were demultiplexed and adaptor-trimmed using Porechop v0.2.4 (Porechop, https://github.com/rrwick/Porechop). The filtered reads were then clustered, polished, and taxonomically classified by NanoCLUST (Rodriguez-Perez et al., 2021) based on the full-length 16S rRNA gene sequences from the Ribosomal Database Project (RDP) database (Cole et al., 2003)”.

HDLc and triglyceride concentrations were measured using the Alinity C (Abbott Laboratories, USA; Otawara-Shi, Tochigi-Ken, Japan) with accelerator selective detergent (HDL cholesterol, HDLc), and glycerol phosphate oxidase (triglyceride) procedures. HDLc and triglyceride coefficients of variation were 2.6%, and 2.2%, respectively. In our study, we used HDLc as well as a z unit-based composite scores reflecting the atherogenic index of plasma (zAIP) as z triglycerides – z HDLc (Morelli et al., 2021; Mousa et al., 2022).

### Statistical analysis

Analysis of variance and univariate GLM analysis were used to determine the differences between study groups regarding continuous variables. At p<0.05, pairwise comparisons of group means were performed to identify differences between the three study groups. In addition, multiple comparisons were corrected using the false discovery rate (FDR) p-value (Benjamini and Hochberg, 1995). Analysis of contingency tables was used to make comparisons between variables based on categories (Chi-square tests). Correlations between variables were examined using Pearson’s product-moment correlation coefficients. While allowing for the effects of sex, age, education, and BMI, multivariate regression analyses were conducted to determine the best predictors of the phenome of depression. In addition to the manual regression method, we also utilized an automated method with p-values of 0.05 for model entry and 0.10 for model elimination. We calculated the model statistics (F, df, and p values) and total variance explained (R^2^), and for each predictor, the standardized beta coefficients with t statistics and exact p-values. In addition, the variance inflation factor and tolerance were assessed to detect any collinearity or multicollinearity issues. Using the White and modified Breusch-Pagan homoscedasticity tests, heteroskedasticity was determined. We have used IBM, SPSS windows version 28 to perform all the above statistical analyses. Moreover, we employed different automatic regression analyses to define the best microbiota phyla, genera, and species data predicting SB, PC_STROOP and the phenome of MDD: a) ridge regression analysis (λ=0.1) with tolerance = 0.4 (using Statistica, windows version 12); a) forward stepwise automatic linear modeling analyses with the overfit criterion as entry/removal criterion with maximum effects number of 6; and c) best subsets with overfit prevention criterion performed on the 20 most important microbiota obtained in regressions a and b (both performed with SPSS 28). Following these analyses we performed manual regression analysis using SPSS 28 and Statistica 12 to check the final models for collinearity and residual distributions and to compute and display partial regression analysis of clinical data on the microbiome taxa. We used logarithmic or rank inverse-normal transformations to normalize the data distribution. The phylum, genus, and species microbiota abundance data were processed in isometric log-ratio (ILR) Box-Cox transformation (ILR abundance), while microbiota data with less than 35% measurable data were entered as dummy variables (prevalence). The significance level of all statistical analyses was determined using 0.05-valued two-tailed tests.

PCA was used as a feature reduction method to construct new PCs that reflect an underlying concept. Towards this end, the variance explained (VE) by the first PC should be at least > 50%, while all variables should show high loadings on the first PC (namely > 0.66).

Furthermore, the factoriability of the correlation matrix was checked with the Kaiser-Meyer-Olkin (KMO) Measure of Sampling Adequacy (values <0.5 indicate that remedial actions should be taken and values > 0.7 indicate a more than adequate sampling). The sphericity test developed by Bartlett is used in order to test the null hypothesis that the variables included in the population correlation matrix are uncorrelated. Moreover, we also inspected the anti-image correlation matrix as an index of sampling adequacy. Two-step clustering analysis was performed to discover whether a valid cluster of MDD patients could be retrieved based on the microbiome and clinical data (number of clusters prespecified as 3, Schwarz’s Bayesian Criterion). The clustering quality was evaluated using the silhouette measure of cohesion and separation, which should be >0.5 (indicating an adequate cluster solution). According to the results of an a priori calculation of the sample size performed with G*Power 3.1.9.4 (multiple regression analysis with 6 covariates), the estimated sample size should be 65 when using an effect size of 0.2 at p=005 (two-tailed) and power = 0.08.

Using partial least squares (PLS) path analysis (SmartPLS) (Ringle et al, 2012; Hair et al., 2019), we investigated the potential causal links between ACE, ROI, the microbiome and the phenome of depression. PLS path analysis was only carried out if both the inner and the outer models satisfied the quality requirements outlined in the following list: a) the overall model fit, namely the standardized root mean square residuals (SRMR) is satisfactory, namely SRMR <0.08; b) the outer latent vectors exhibit accurate construct and convergence validity, as shown by average variance extracted (AVE) > 0.5, composite reliability > 0.8, rho A > 0.8, Cronbach’s alpha > 0.7; and all outer loadings > 0.66 at p <0.001, d) the model’s prediction performance is adequate using PLSPredict, and e) Confirmatory Tetrad Analysis (CTA) shows that the outer models are not mis-specified as reflective models. In the event that all of the aforementioned model quality data satisfy the predetermined criteria, we carry out a complete PLS path analysis with 5,000 bootstrap samples, produce the path coefficients (with exact p-values), and additionally compute the specific and total indirect (that is, mediated) effects in addition to the total effects.

## Results

### Results of PCA

We were able to extract reliable PCs from the 4 ACE items denoting sexual abuse (labeled PC_sexabuse) (KMO=0.565, Bartlett’s sphericity test χ2=126.547, df=6, p<0.001, VE=59.00%, all loadings >0.646) and 5 ACE items denoting emotional neglect (labeled PC_ emneglect) (KMO=0.869, Bartlett’s sphericity test χ2=409.137, df=10, p<0.001, VE=86.82%, all loadings >0.919). Since we were unable to extract one PC from the 5 ACE physical neglect data, we used the z composite score of the sum of the 5 items in the analyses (dubbed: Comp_physneglect). We also extracted PCs from the physical and emotional abuse items (dubbed as PC_physabuse and PC_emabuse, respectively). We were able to extract one reliable PC from PC_sexabuse, PC_emabuse and PC_physabuse (labeled PC_abuse) (KMO=0.565, Bartlett’s spheriticy test χ2=25.014, df=3, p<0.001, VE=56.08%, all loadings >0.676). Overall neglect was conceptualized as the first PC extracted from PC_emneglect and Comp_physneglect scores (labeled as PC_neglect). We were able to extract one PC from the three Stroop subtest scores (KMO=0.572, Bartlett’s sphericity test χ2=60.108, df=3, p<0.001, VE=65.75%, all loadings >0.719), labeled PC_Stroop.

Table 1 shows that we were able to extract one reliable PC from the total number of episodes, LT_SI and LT_SA (labeled ROI); and one reliable PC from the total BDI and HAMD scores and current SBs (labeled PC_phenome). We were also able to extract one reliable PC from the number of depressive episodes, LT_SB, total BDI and HAMD scores, and Curr_SB (labeled PC_ROI+phenome).

**Table 1.** Results of principal component analysis (PCA)

| PCA#1: ROI |  | PCA#2: Phenome |  | PCA#3: ROI+phenome |  | PCA#4: Enterotype1+ROI |  | PCA#5: Enterotype1+ROI-phenome |  | PCA#6: LT traject+Enterotype1 |  |
| --- | --- | --- | --- | --- | --- | --- | --- | --- | --- | --- | --- |
| Variables | Loadings | variables | Loadings | Variables | Loadings | Variables | Loadings | Variables | Loadings | Variables | Loadings |
| # episodes | 0.840 | Curr_SB | 0.836 | # episodes | 0.867 | # episodes | 0.839 | ROI | 0.886 | PC_Abuse | 0.702 |
| LT_SI | 0.939 | BDI | 0.887 | LT_SB | 0.851 | LT_SI | 0.909 | BDI | 0.823 | ROI | 0.883 |
| LT_SA | 0.826 | HAMD | 0.907 | BDI | 0.843 | LT_SA | 0.790 | HAMD | 0.880 | Phenome | 0.929 |
|  |  |  |  | HAMD | 0.902 | Enterotype1 | 0.691 | Curr_SB | 0.812 | Enterotype1 | 0.695 |
|  |  |  |  | Curr_SB | 0.820 |  |  | Enterotype1 | 0.694 |  |  |
| KMO=0.614 |  | KMO=0.708 |  | KMO=0.821 |  | KMO=0.693 |  | KMO=0.843 |  | KMO=0.719 |  |
| X<sup>2</sup>=92.769 (df=3)<br>p<0.001 |  | X<sup>2</sup>=90.029 (df=3)<br>p<0.001 |  | X<sup>2</sup>=228.327 (df=10)<br>p<0.001 |  | X<sup>2</sup>=112.714 (df=6)<br>p<0.001 |  | X<sup>2</sup>=152.931 (df=10)<br>p<0.001 |  | X<sup>2</sup>=105.052 (df=6)<br>p<0.001 |  |
| VE=75.65% |  | VE=76.85% |  | VE=73.48% |  | VE=65.769% |  | VE=67.54% |  | VE=65.48% |  |

LT\_SI: lifetime suicidal ideation, LT\_SA: lifetime suicidal attempts, Curr\_SB: current suicidal behaviors, BDI: Beck Depression Inventory score; HAMD: Hamilton Depression Rating Scale score; ROI: recurrence of illness index; PC\_abuse: index of physical + emotional + sexual abuse; Enterotype 1 and 2: two dysbiosis indices, the first reflecting depression and the second antioxidant-metabolic alterations of depression; LT\_traject: lifetime trajectory, namely including childhood abuse data. KMO: Kaiser-Meyer-Olkin measure of sampling adequacy; X<sup>2</sup>: results of Bartlett's test of sphericity; VE: variance explained by the first PC.

### Construction of the first enterotype

**Table 2** shows the outcome of two different multiple regression analyses with PC_phenome as the dependent variable and the microbiome taxa as explanatory variables. Both linear modeling analysis and ridge regression analysis showed basically the same results. Using linear modeling analysis, up to 36.1% of the variance in PC_phenome was explained by 6 taxa, namely *Hungatella* and *Fusicatenibacter (*both positively) and *Butyricicoccus, Clostridium, Parabacteroides merdae,* and *Desulfovibrio piger* (all inversely associated). **Figure 1** shows the partial regression of PC_phenome on *Butyricicoccus.* Ridge regression showed that 34.3% of the variance in PC_phenome was explained by the same taxa, except *P. merdae.* Consequently, we have computed a z unit-based composite score (labeled enterotype 1) based on the sum of the 6 taxa with z transformation of *z Fusicatenibacter + Hungatella* (0 or 1 score) *-* z *Butyricicoccus -* z *Clostridium -* z *P. merdae – D. piger* (0 or 1 score).

**Figure 1.**
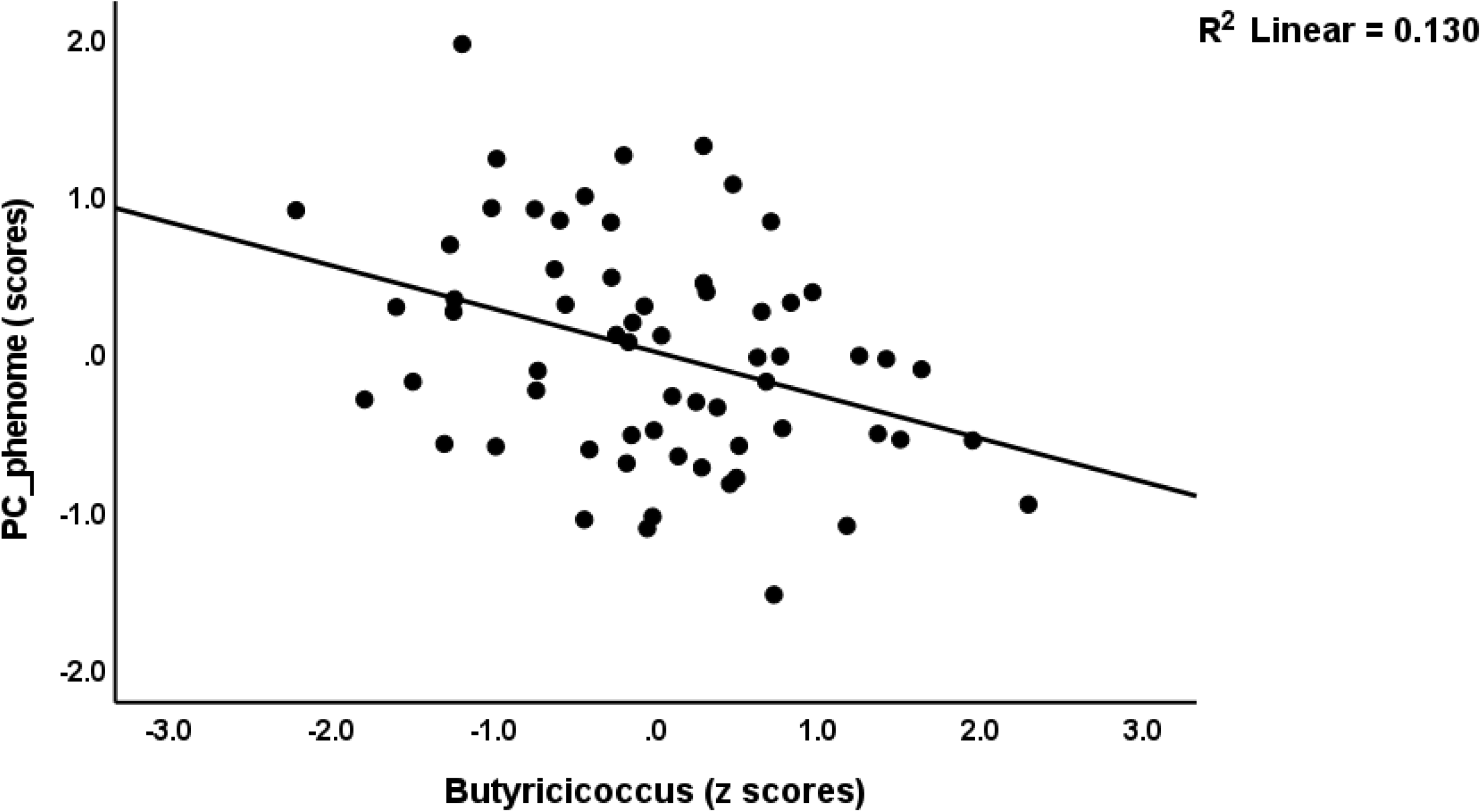
Partial regression of the phenome of depression (PC_phenome) on the isometric log-ratio abundance of *Butyricicoccus*

**Table 2.**
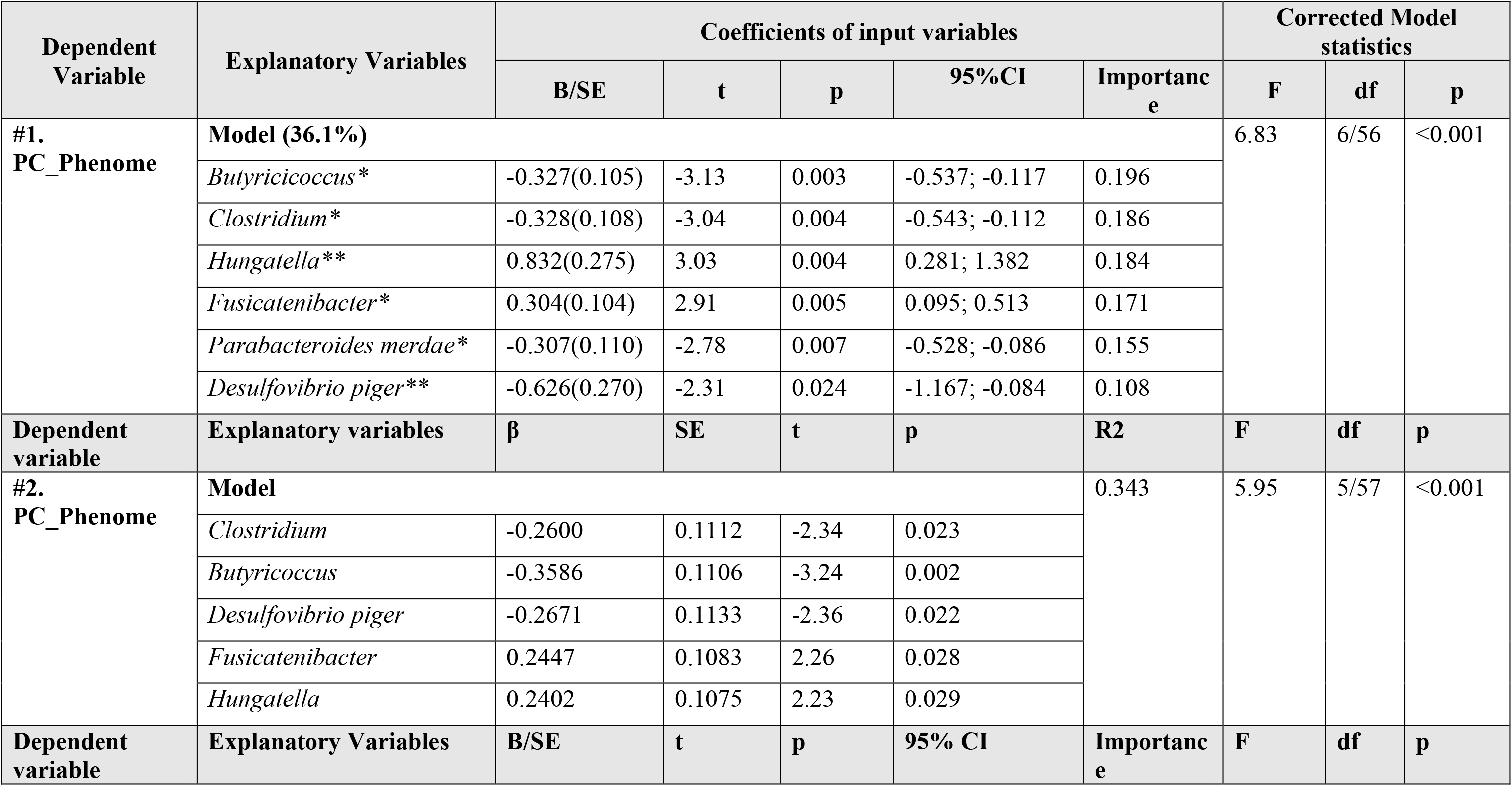
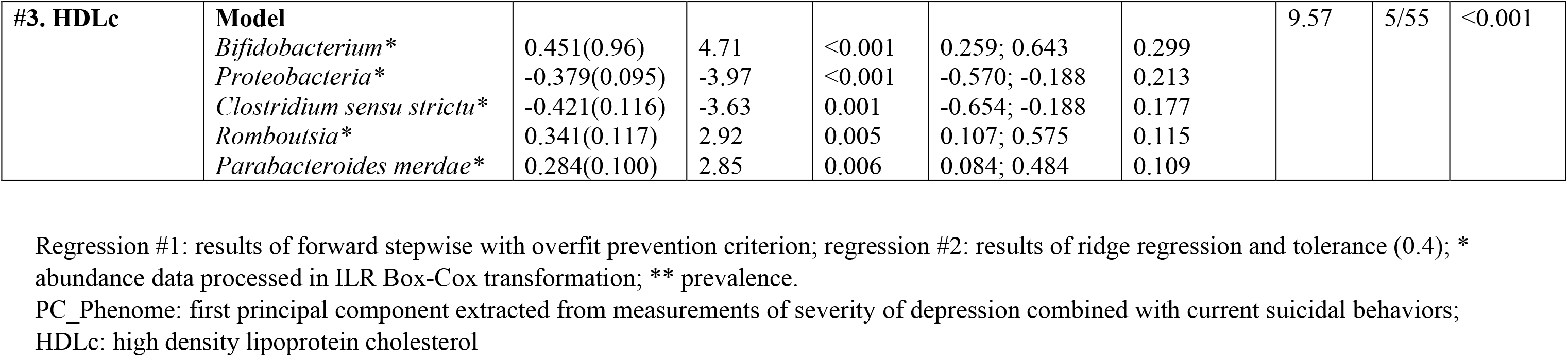
Results of linear modelling analyses with the phenome scores as dependent variables, and microbiota assessments as explanatory variables, while allowing for the effects of age, sex, body mass index, and drug status.

| Dependent Variable | Explanatory Variables | Coefficients of input variables |  |  |  |  | Corrected Model statistics |  |  |
| --- | --- | --- | --- | --- | --- | --- | --- | --- | --- |
|  |  | B/SE | t | p | 95%CI | Importance | F | df | p |
| #1.<br>PC_Phenome | **Model (36.1%)** |  |  |  |  |  | 6.83 | 6/56 | <0.001 |
|  | *Butyricicoccus* * | -0.327(0.105) | -3.13 | 0.003 | -0.537; -0.117 | 0.196 |  |  |  |
|  | *Clostridium* * | -0.328(0.108) | -3.04 | 0.004 | -0.543; -0.112 | 0.186 |  |  |  |
|  | *Hungatella* ** | 0.832(0.275) | 3.03 | 0.004 | 0.281; 1.382 | 0.184 |  |  |  |
|  | *Fusicatenibacter* * | 0.304(0.104) | 2.91 | 0.005 | 0.095; 0.513 | 0.171 |  |  |  |
|  | *Parabacteroides merdae* * | -0.307(0.110) | -2.78 | 0.007 | -0.528; -0.086 | 0.155 |  |  |  |
|  | *Desulfovibrio piger* ** | -0.626(0.270) | -2.31 | 0.024 | -1.167; -0.084 | 0.108 |  |  |  |
| Dependent variable | Explanatory variables | $\beta$ | SE | t | p | R2 | F | df | p |
| #2.<br>PC_Phenome | **Model** |  |  |  |  | 0.343 | 5.95 | 5/57 | <0.001 |
|  | *Clostridium* | -0.2600 | 0.1112 | -2.34 | 0.023 |  |  |  |  |
|  | *Butyricoccus* | -0.3586 | 0.1106 | -3.24 | 0.002 |  |  |  |  |
|  | *Desulfovibrio piger* | -0.2671 | 0.1133 | -2.36 | 0.022 |  |  |  |  |
|  | *Fusicatenibacter* | 0.2447 | 0.1083 | 2.26 | 0.028 |  |  |  |  |
|  | *Hungatella* | 0.2402 | 0.1075 | 2.23 | 0.029 |  |  |  |  |
| Dependent variable | Explanatory Variables | B/SE | t | p | 95% CI | Importance | F | df | p |
| #3. HDLc | **Model** |  |  |  |  |  | 9.57 | 5/55 | <0.001 |
|  | *Bifidobacterium* * | 0.451(0.96) | 4.71 | <0.001 | 0.259; 0.643 | 0.299 |  |  |  |
|  | *Proteobacteria* * | -0.379(0.095) | -3.97 | <0.001 | -0.570; -0.188 | 0.213 |  |  |  |
|  | *Clostridium sensu strictu* * | -0.421(0.116) | -3.63 | 0.001 | -0.654; -0.188 | 0.177 |  |  |  |
|  | *Romboutsia* * | 0.341(0.117) | 2.92 | 0.005 | 0.107; 0.575 | 0.115 |  |  |  |
|  | *Parabacteroides merdae* * | 0.284(0.100) | 2.85 | 0.006 | 0.084; 0.484 | 0.109 |  |  |  |

Regression #1: results of forward stepwise with overfit prevention criterion; regression #2: results of ridge regression and tolerance (0.4); \* abundance data processed in ILR Box-Cox transformation; \*\* prevalence.
PC\_Phenome: first principal component extracted from measurements of severity of depression combined with current suicidal behaviors;
HDLc: high density lipoprotein cholesterol

Table 1 shows that one reliable PC could be extracted from the three 3 ROI indices and enterotype 1 (PCA #4), indicating that the latter is strongly associated with ROI. Moreover, one validated PC (PCA#5) could be extracted from ROI, BDI, HAMD, Curr_SB and enterotype 1, indicating that the latter belongs to the same core as the ROI-phenome association. Finally, we were also able to extract one PC from PC_abuse, ROI, PC_phenome and enterotype1 (PCA#6).

Consequently, we have examined whether we could retrieve a more severe MDD class and, therefore, performed clustering analysis with diagnosis, ROI, enterotype 1, and PC_phenome data as clustering variables. Table 3 shows that three clusters were formed, namely healthy controls (n=37), MDD patients with less severe features (labeled simple dysmood disorder or SDMD, n=17) and those with more severe features (labeled major dysmood disorder or MDMD, n=12). Of course, we did not carry out this analysis to define diagnostic criteria for both clusters as this would need a larger study group and cross-validation. The only aim is to show the demographic, clinical and biomarkers data measurements in controls versus patients divided into less and more severe patients. It should be stressed that the primary outcome data of this study are the multiple regression (including PLS) analyses.

**Table 3:**
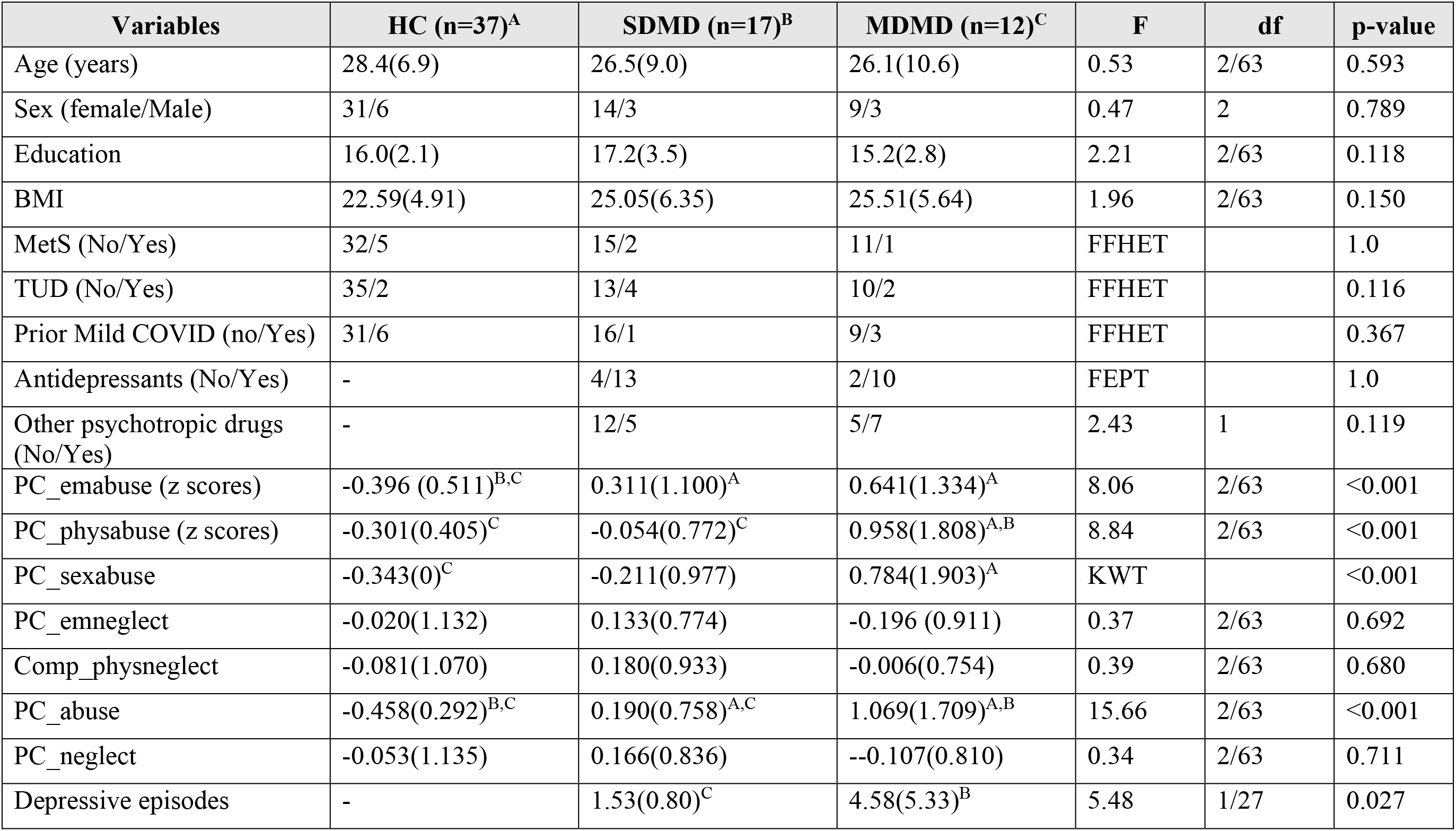
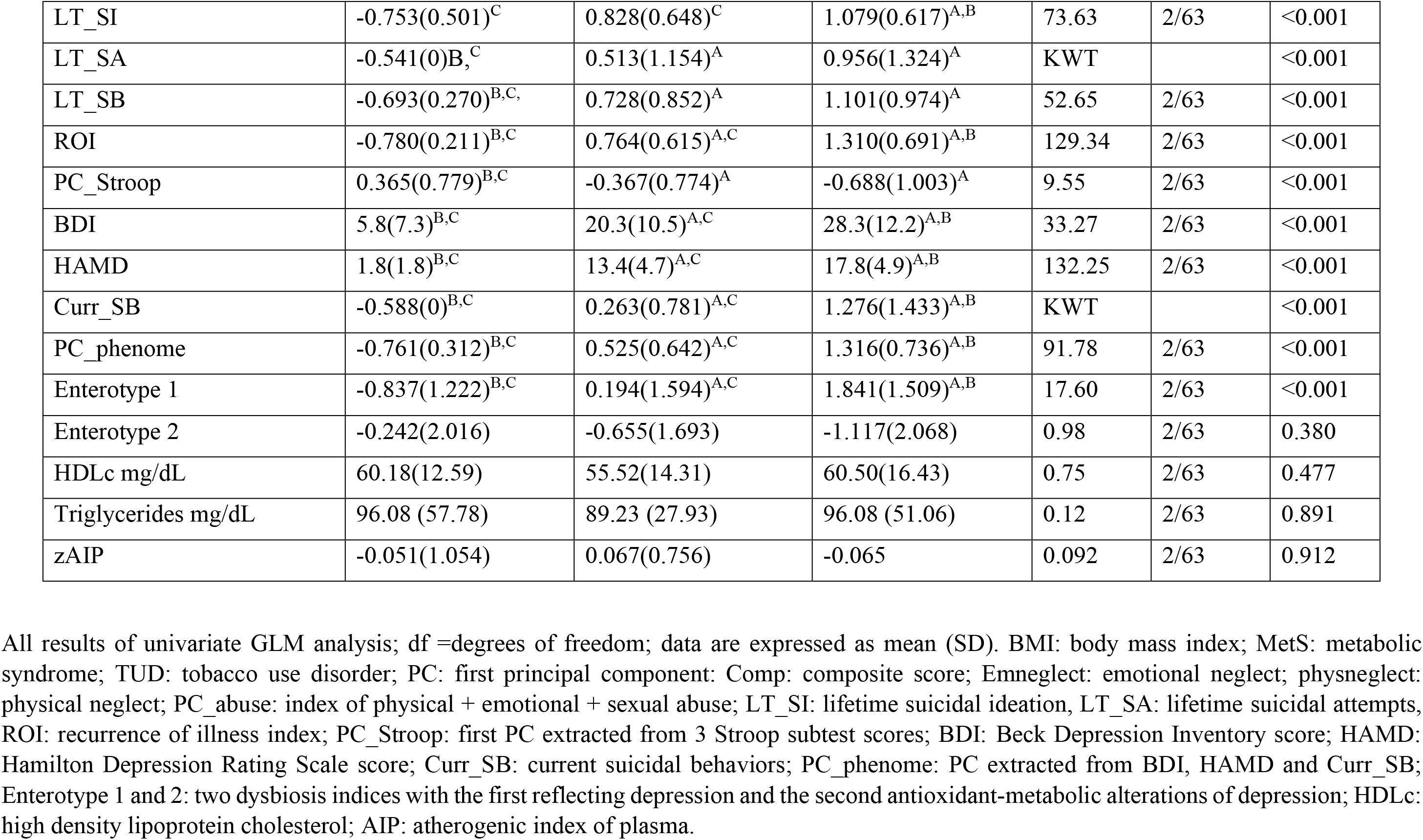
Sociodemographic, clinical and biomarker data in healthy controls (HC) and major depressed (MDD) patients divided into those with major (MDMD) and simple (SDMD) dysmood disorder

| Variables | HC (n=37)<sup>A</sup> | SDMD (n=17)<sup>B</sup> | MDMD (n=12)<sup>C</sup> | F | df | p-value |
| --- | --- | --- | --- | --- | --- | --- |
| Age (years) | 28.4(6.9) | 26.5(9.0) | 26.1(10.6) | 0.53 | 2/63 | 0.593 |
| Sex (female/Male) | 31/6 | 14/3 | 9/3 | 0.47 | 2 | 0.789 |
| Education | 16.0(2.1) | 17.2(3.5) | 15.2(2.8) | 2.21 | 2/63 | 0.118 |
| BMI | 22.59(4.91) | 25.05(6.35) | 25.51(5.64) | 1.96 | 2/63 | 0.150 |
| MetS (No/Yes) | 32/5 | 15/2 | 11/1 | FFHET |  | 1.0 |
| TUD (No/Yes) | 35/2 | 13/4 | 10/2 | FFHET |  | 0.116 |
| Prior Mild COVID (no/Yes) | 31/6 | 16/1 | 9/3 | FFHET |  | 0.367 |
| Antidepressants (No/Yes) | - | 4/13 | 2/10 | FEPT |  | 1.0 |
| Other psychotropic drugs (No/Yes) | - | 12/5 | 5/7 | 2.43 | 1 | 0.119 |
| PC_emabuse (z scores) | -0.396 (0.511)<sup>B,C</sup> | 0.311(1.100)<sup>A</sup> | 0.641(1.334)<sup>A</sup> | 8.06 | 2/63 | <0.001 |
| PC_physabuse (z scores) | -0.301(0.405)<sup>C</sup> | -0.054(0.772)<sup>C</sup> | 0.958(1.808)<sup>A,B</sup> | 8.84 | 2/63 | <0.001 |
| PC_sexabuse | -0.343(0)<sup>C</sup> | -0.211(0.977) | 0.784(1.903)<sup>A</sup> | KWT |  | <0.001 |
| PC_emneglect | -0.020(1.132) | 0.133(0.774) | -0.196 (0.911) | 0.37 | 2/63 | 0.692 |
| Comp_physneglect | -0.081(1.070) | 0.180(0.933) | -0.006(0.754) | 0.39 | 2/63 | 0.680 |
| PC_abuse | -0.458(0.292)<sup>B,C</sup> | 0.190(0.758)<sup>A,C</sup> | 1.069(1.709)<sup>A,B</sup> | 15.66 | 2/63 | <0.001 |
| PC_neglect | -0.053(1.135) | 0.166(0.836) | --0.107(0.810) | 0.34 | 2/63 | 0.711 |
| Depressive episodes | - | 1.53(0.80)<sup>C</sup> | 4.58(5.33)<sup>B</sup> | 5.48 | 1/27 | 0.027 |
| LT_SI | -0.753(0.501)<sup>C</sup> | 0.828(0.648)<sup>C</sup> | 1.079(0.617)<sup>A,B</sup> | 73.63 | 2/63 | <0.001 |
| LT_SA | -0.541(0)<sup>B,C</sup> | 0.513(1.154)<sup>A</sup> | 0.956(1.324)<sup>A</sup> | KWT |  | <0.001 |
| LT_SB | -0.693(0.270)<sup>B,C</sup> | 0.728(0.852)<sup>A</sup> | 1.101(0.974)<sup>A</sup> | 52.65 | 2/63 | <0.001 |
| ROI | -0.780(0.211)<sup>B,C</sup> | 0.764(0.615)<sup>A,C</sup> | 1.310(0.691)<sup>A,B</sup> | 129.34 | 2/63 | <0.001 |
| PC_Stroop | 0.365(0.779)<sup>B,C</sup> | -0.367(0.774)<sup>A</sup> | -0.688(1.003)<sup>A</sup> | 9.55 | 2/63 | <0.001 |
| BDI | 5.8(7.3)<sup>B,C</sup> | 20.3(10.5)<sup>A,C</sup> | 28.3(12.2)<sup>A,B</sup> | 33.27 | 2/63 | <0.001 |
| HAMD | 1.8(1.8)<sup>B,C</sup> | 13.4(4.7)<sup>A,C</sup> | 17.8(4.9)<sup>A,B</sup> | 132.25 | 2/63 | <0.001 |
| Curr_SB | -0.588(0)<sup>B,C</sup> | 0.263(0.781)<sup>A,C</sup> | 1.276(1.433)<sup>A,B</sup> | KWT |  | <0.001 |
| PC_phenome | -0.761(0.312)<sup>B,C</sup> | 0.525(0.642)<sup>A,C</sup> | 1.316(0.736)<sup>A,B</sup> | 91.78 | 2/63 | <0.001 |
| Enterotype 1 | -0.837(1.222)<sup>B,C</sup> | 0.194(1.594)<sup>A,C</sup> | 1.841(1.509)<sup>A,B</sup> | 17.60 | 2/63 | <0.001 |
| Enterotype 2 | -0.242(2.016) | -0.655(1.693) | -1.117(2.068) | 0.98 | 2/63 | 0.380 |
| HDLc mg/dL | 60.18(12.59) | 55.52(14.31) | 60.50(16.43) | 0.75 | 2/63 | 0.477 |
| Triglycerides mg/dL | 96.08 (57.78) | 89.23 (27.93) | 96.08 (51.06) | 0.12 | 2/63 | 0.891 |
| zAIP | -0.051(1.054) | 0.067(0.756) | -0.065 | 0.092 | 2/63 | 0.912 |

All results of univariate GLM analysis; df =degrees of freedom; data are expressed as mean (SD). BMI: body mass index; MetS: metabolic syndrome; TUD: tobacco use disorder; PC: first principal component; Comp: composite score; Emneglect: emotional neglect; physneglect: physical neglect; PC\_abuse: index of physical + emotional + sexual abuse; LT\_SI: lifetime suicidal ideation, LT\_SA: lifetime suicidal attempts, ROI: recurrence of illness index; PC\_Stroop: first PC extracted from 3 Stroop subtest scores; BDI: Beck Depression Inventory score; HAMD: Hamilton Depression Rating Scale score; Curr\_SB: current suicidal behaviors; PC\_phenome: PC extracted from BDI, HAMD and Curr\_SB; Enterotype 1 and 2: two dysbiosis indices with the first reflecting depression and the second antioxidant-metabolic alterations of depression; HDLc: high density lipoprotein cholesterol; AIP: atherogenic index of plasma.

### Features of MDD, SDMD and MDMD

**Table 3** shows the socio-demographic, clinical and biomarker data measurements in controls and patients with MDMD and SDMD. There were no significant differences in age, sex, education, BMI, MetS, TUD, and prior mild COVID-19 infection between the three groups. There were no differences in the drug state (use of antidepressants or other psychotropic drugs, namely, atypical antipsychotics: n=4, mood stabilizers: n=1, benzodiazepines: n=8) between MDMD and SDMD. PC_emabuse was significantly greater in patients than in controls. PC_physabuse was significantly higher in MDMD than in the two other groups, whereas PC_sexabuse was greater in MDMD than controls. PC_abuse was significantly different between the three groups and increased from controls to SDMD to MDMD. The number of depressive episodes was significantly higher in MDMD than in SDMD. LT_SB and LT_SA were significantly higher in patients than in controls, while LT_SI was higher in MDMD than in controls and SDMD. ROI was significantly higher in MDMD than in SDMD. Patients with MDD showed significantly lower PC_Stroop values than controls. The total BDI, HAMD, Curr_SB and PC_phenome scores increased from controls → SDMD → MDMD. All differences among these phenome data remained significant after FDR p-correction. The enterotype 1 score was significantly different between the three study groups and increased from controls → SDMD → MDMD. There were no significant differences in HDLc and AIP between the three study groups. Univariate GLM analysis showed no associations between the use of antidepressants and other psychotropic drugs and mild COVID- 19 some months earlier and any of the microbiota and clinical data (even without FDR p-correction).

### Enterotype 1 and clinical features of MDD

**Table 4** shows that enterotype 1 is associated with PC_abuse (but not PC_neglect), ROI and its components (number of episodes and LT_SB), PC_phenome (bot not PC_Stroop), and Current_SB. **Table 5**, regression #1 shows that 38.9% of the variance in enterotype 1 was explained by ROI and male sex (both positively correlated). **Figure 2** shows the partial regression of enterotype 1 on ROI. Removal of ROI from this analysis (regression #2) shows that PC_abuse and male sex explained 19.6% of the variance in enterotype 1. A large part of the variance (70.2%) in PC_phenome (regression #3) was explained by ROI, PC_abuse and enterotype 1. Deleting ROI from this analysis (regression #4) showed that 56.2% of the variance in PC_phenome was explained by enterotype 1, PC_emabuse and PC_sexabuse (all positively associated).

**Figure 2.**
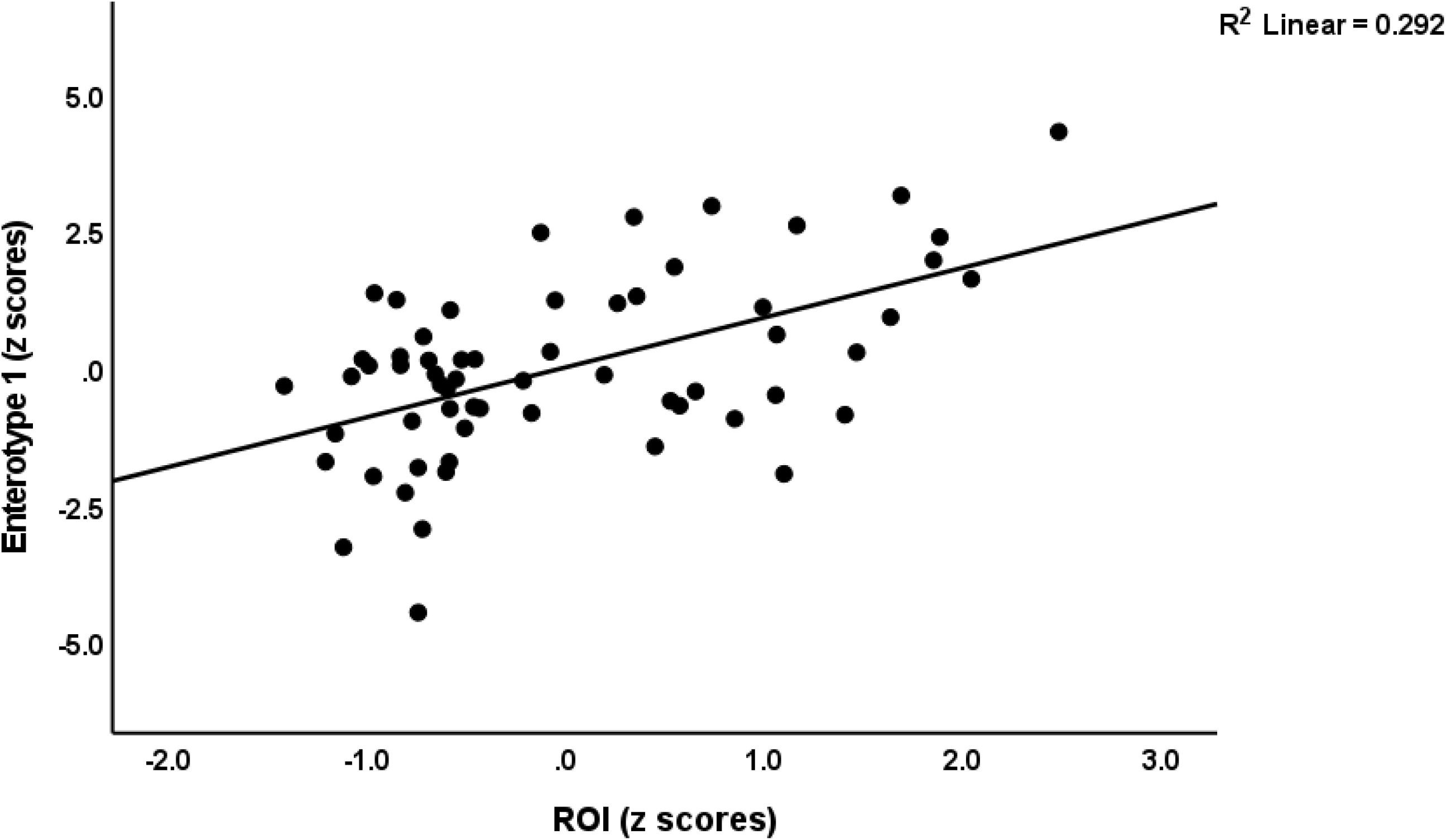
Partial regression of enterotype 1, a gut dysbiosis index, on the recurrence of illness index (ROI)

**Table 4.** Intercorrelation matrix

| **Variables** | **Enterotype 1** | **PC abuse** | **#Episodes** | **LT_SB** | **ROI** | **PC_Stroop** | **HDLc** |
| --- | --- | --- | --- | --- | --- | --- | --- |
| **PC_abuse** | 0.340 (0.006) |  | 0.426 (<0.001) | 0.534 (<0.001) | 0.544 (<0.001) | -0.315 (0.008) | -0.296 (0.014) |
| **PC_neglect** | -0.048 (0.709) | -0.191 (0.115) | -0.007 (0.956) | 0.035 (0.782) | 0.029 (0.819) | 0.014 (0.911) | 0.128 (0.297) |
| **# episodes** | 0.526 (<0.001) | 0.461 (<0.001) | - | 0.593 (<0.001) | 0.844 (<0.001) | -0.306 (0.011) | -0.131 (0.288) |
| **LT_SB** | 0.471 (<0.001) | 0.534 (<0.001) | 0.593 (<0.001) | - | 0.952 (<0.001) | -0.386 (0.001) | -0.146 (0.243) |
| **ROI** | 0.531 (<0.001) | 0.544 (<0.001) | 0.810 (<0.001) | 0.952 (<0.001) | - | -0.436 (<0.001) | -0.129 (0.301) |
| **PC_Stroop** | -0.105 (0.414) | -0.315 (0.008) | -0.306 (0.011) | -0.386 (0.001) | -0.436 (<0.001) | - | 0.091 (0.458) |
| **PC_phenome** | 0.599 (<0.001) | 0.569 (<0.001) | 0.743 (<0.001) | 0.746 (<0.001) | 0.822 (<0.001) | -0.381 (0.001) | -0.298 (0.013) |
| **Current_SB** | 0.524 (<0.001) | 0.586 (<0.001) | 0.709 (<0.001) | 0.755 (<0.001) | 0.744 (<0.001) | -0.347 (0.003) | -0.273 (0.024) |
| **Enterotype 2** | -0.012 (0.929) | -0.275 (0.029) | -0.152 (0.212) | -0.162 (0.190) | -0.183 (0.157) | 0.168 (0.167) | 0.696 (<0.001) |

PC\_abuse: first principal component (PC) extracted from physical + emotional + sexual abuse; PC\_neglect: PC extracted from childhood neglect scores; #episodes: number of depressive episodes; LT\_SB: lifetime suicidal behaviors; ROI: recurrence of illness index; PC\_Stroop: PC extracted from 3 Stroop test results; PC\_phenome: PC extracted from severity of depression and Curr\_SB (current suicidal behaviors); enterotype 1 and 2: two dysbiosis indices with the first reflecting depression and the second antioxidant-metabolic alterations of depression; HDLc: high density lipoprotein

**Table 5.**
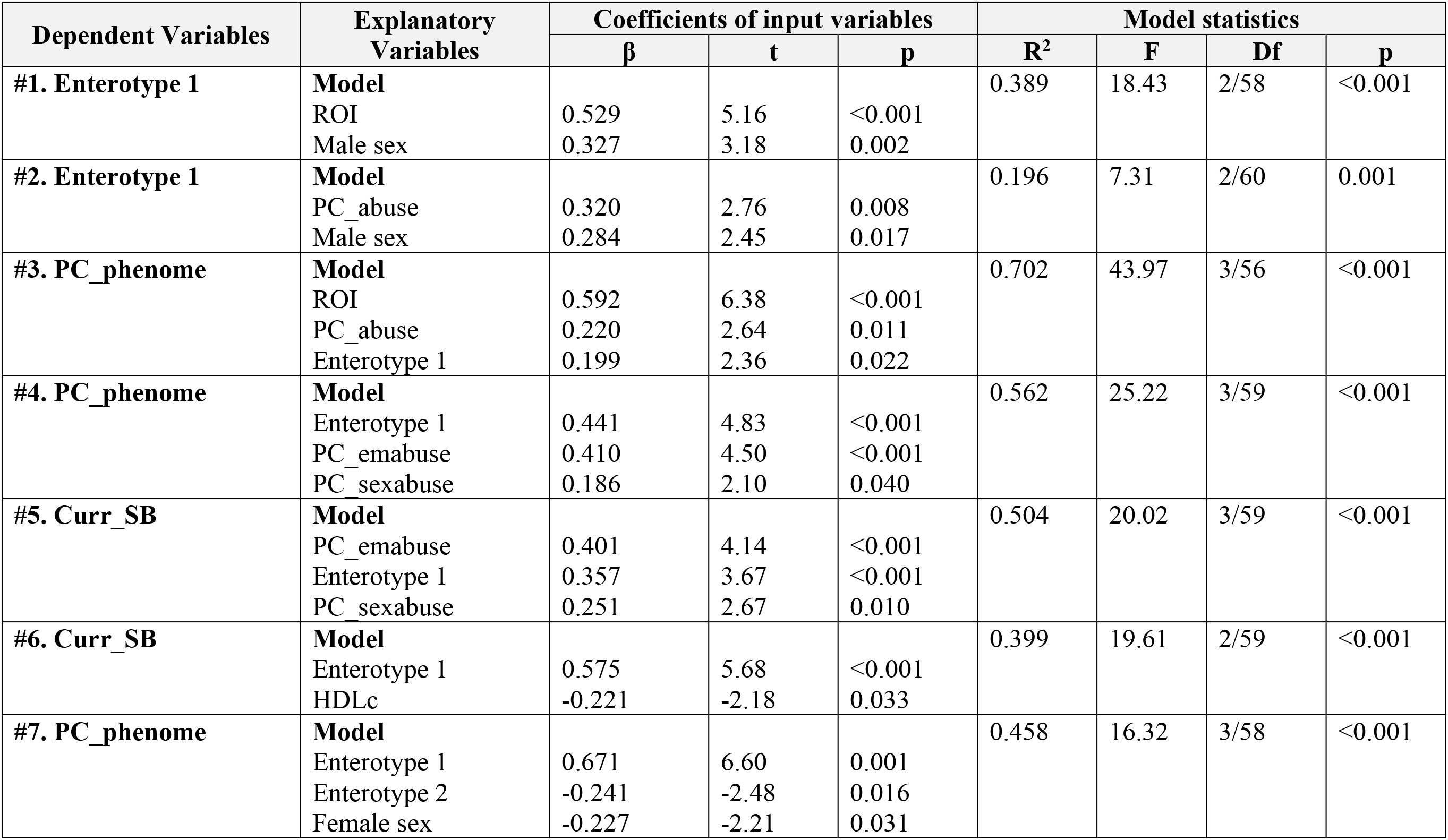
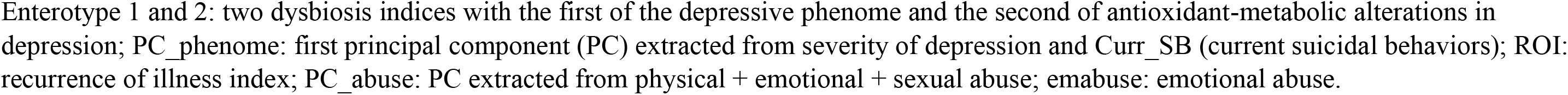
Results of multiple regression analyses with phenome or microbiota set scores as dependent variables.

| Dependent Variables | Explanatory Variables | Coefficients of input variables |  |  | Model statistics |  |  |  |
| --- | --- | --- | --- | --- | --- | --- | --- | --- |
| | | $\beta$ | t | p | R<sup>2</sup> | F | Df | p |
| #1. Enterotype 1 | **Model** |  |  |  | 0.389 | 18.43 | 2/58 | <0.001 |
|  | ROI | 0.529 | 5.16 | <0.001 |  |  |  |  |
|  | Male sex | 0.327 | 3.18 | 0.002 |  |  |  |  |
| #2. Enterotype 1 | **Model** |  |  |  | 0.196 | 7.31 | 2/60 | 0.001 |
|  | PC_abuse | 0.320 | 2.76 | 0.008 |  |  |  |  |
|  | Male sex | 0.284 | 2.45 | 0.017 |  |  |  |  |
| #3. PC_phenome | **Model** |  |  |  | 0.702 | 43.97 | 3/56 | <0.001 |
|  | ROI | 0.592 | 6.38 | <0.001 |  |  |  |  |
|  | PC_abuse | 0.220 | 2.64 | 0.011 |  |  |  |  |
|  | Enterotype 1 | 0.199 | 2.36 | 0.022 |  |  |  |  |
| #4. PC_phenome | **Model** |  |  |  | 0.562 | 25.22 | 3/59 | <0.001 |
|  | Enterotype 1 | 0.441 | 4.83 | <0.001 |  |  |  |  |
|  | PC_emabuse | 0.410 | 4.50 | <0.001 |  |  |  |  |
|  | PC_sexabuse | 0.186 | 2.10 | 0.040 |  |  |  |  |
| #5. Curr_SB | **Model** |  |  |  | 0.504 | 20.02 | 3/59 | <0.001 |
|  | PC_emabuse | 0.401 | 4.14 | <0.001 |  |  |  |  |
|  | Enterotype 1 | 0.357 | 3.67 | <0.001 |  |  |  |  |
|  | PC_sexabuse | 0.251 | 2.67 | 0.010 |  |  |  |  |
| #6. Curr_SB | **Model** |  |  |  | 0.399 | 19.61 | 2/59 | <0.001 |
|  | Enterotype 1 | 0.575 | 5.68 | <0.001 |  |  |  |  |
|  | HDLc | -0.221 | -2.18 | 0.033 |  |  |  |  |
| #7. PC_phenome | **Model** |  |  |  | 0.458 | 16.32 | 3/58 | <0.001 |
|  | Enterotype 1 | 0.671 | 6.60 | 0.001 |  |  |  |  |
|  | Enterotype 2 | -0.241 | -2.48 | 0.016 |  |  |  |  |
|  | Female sex | -0.227 | -2.21 | 0.031 |  |  |  |  |

Enterotype 1 and 2: two dysbiosis indices with the first of the depressive phenome and the second of antioxidant-metabolic alterations in depression; PC\_phenome: first principal component (PC) extracted from severity of depression and Curr\_SB (current suicidal behaviors); ROI: recurrence of illness index; PC\_abuse: PC extracted from physical + emotional + sexual abuse; emabuse: emotional abuse.

Table 5, regression #5 shows that 50.4% f the variance in Curr_SB is explained by PC_emabuse, enterotype 1, and sex. In order to further explore the associations between SB and enterotype 1, we have carried out PCA and were able to extract one reliable PC from enterotype 1 (loading=0.760). LT_SB (0.877) and current_SB (0.798) (KMO=0.663, Bartlett’s sphericity test χ2=62.421, df=3, p<0.001, VE=71.51%). In addition, 39.9% of the variance in Curr_SB (regression #6) could be explained by the regression on enterotype 1 (positive) and HDLc (inversely).

**Table 6** shows the results of forward stepwise regressions with overfit prevention criterion on microbiome taxa. Overall, SB (regression #1) was best predicted by Enterotype 1 and *Proteobacteria* (both positively associated). PC_Stroop (regression #2) was best predicted by *Intestinimonas* (positively) *and Dialister* (inversely) abundances.

**Table 6:** Results of linear modeling with overfit prevention criterion with clinical data as dependent variables and microbiota as explanatory variables, while allowing for the effects of age, sex, body mass index, and drug status

| Dependent Variables | Explanatory Variables | Coefficients of input variables |  |  |  |  | Corrected Model statistics |  |  |
| --- | --- | --- | --- | --- | --- | --- | --- | --- | --- |
|  |  | B(SE) | t | p | 95%CI | Importance | F | df | p |
| #1. Overall SB | **Model** |  |  |  |  |  | 8.99 | 4/58 | <0.001 |
|  | Enterotype 1 | 0.288(0.059) | 4.93 | <0.001 | 0.171;0.406 | 0.685 |  |  |  |
|  | *Proteobacteria* * | 0.221(0.105) | 2.11 | 0.039 | 0.012; 0.431 | 0.126 |  |  |  |
| #2. PC_Stroop | **Model** |  |  |  |  |  | 4.12 | 6/56 | 0.002 |
|  | *Intestinimonas* * | 0.382(0.132) | 2.89 | 0.005 | 0.117;0.648 | 0.288 |  |  |  |
|  | *Dialister* * | -0.288(0.119) | -2.42 | 0.019 | -0.527; -0.050 | 0.202 |  |  |  |
| #3. AIP | **Model** |  |  |  |  |  | 9.23 | 6/55 | <0.001 |
|  | Female sex | -1.46(0.260) | -4.02 | <0.001 | -1.568; -0.525 | 0.291 |  |  |  |
|  | *Bifidobacterium* * | -0.309(0.097) | -3.17 | 0.003 | -0.504; -0.113 | 0.180 |  |  |  |
|  | *Acidaminococcus* ** | 0.722(0.234) | 3.09 | 0.003 | 0.254; 1.190 | 0.172 |  |  |  |
|  | *Sutterella* * | 0.278(0.094) | 2.97 | 0.004 | 0.090; 0.466 | 0.158 |  |  |  |
|  | *Clostridium sensu strictu* * | 0.254(0.094) | 2.69 | 0.009 | 0.065; 0.442 | 0.130 |  |  |  |
|  | *Verrucomicrobia* * | -0.185(0.094) | -1.96 | 0.055 | -0.374; 0.004 | 0.069 |  |  |  |

\* abundance data processed in ILR Box-Cox transformation; \*\* prevalence
Enterotype 1: a dysbiosis index of depression; Overall SB: first principal component extracted from lifetime and current suicidal behaviors; PC\_Stroop: first principal component extracted from 3 Stroop tests results; AIP: atherogenic index of plasma.

### Enterotype of atherogenicity in MDD

Table 2, regression #3, shows the results of a forward stepwise analysis with overfit prevention criterion. We found that *Proteobacteria* and *Clostridium sensu stricto* abundances were significantly and inversely associated with HDLc, and that *Bifidobacterium, Romboutsia* and *P. merdae* were positively associated. Consequently, we built a z unit-based composite score based on those 5 microbiota taxa, dubbed enterotype 2. **Figure 3** shows the partial regression of HDLc on the ILR abundance of *Bifidobacterium*.

**Figure 3.**
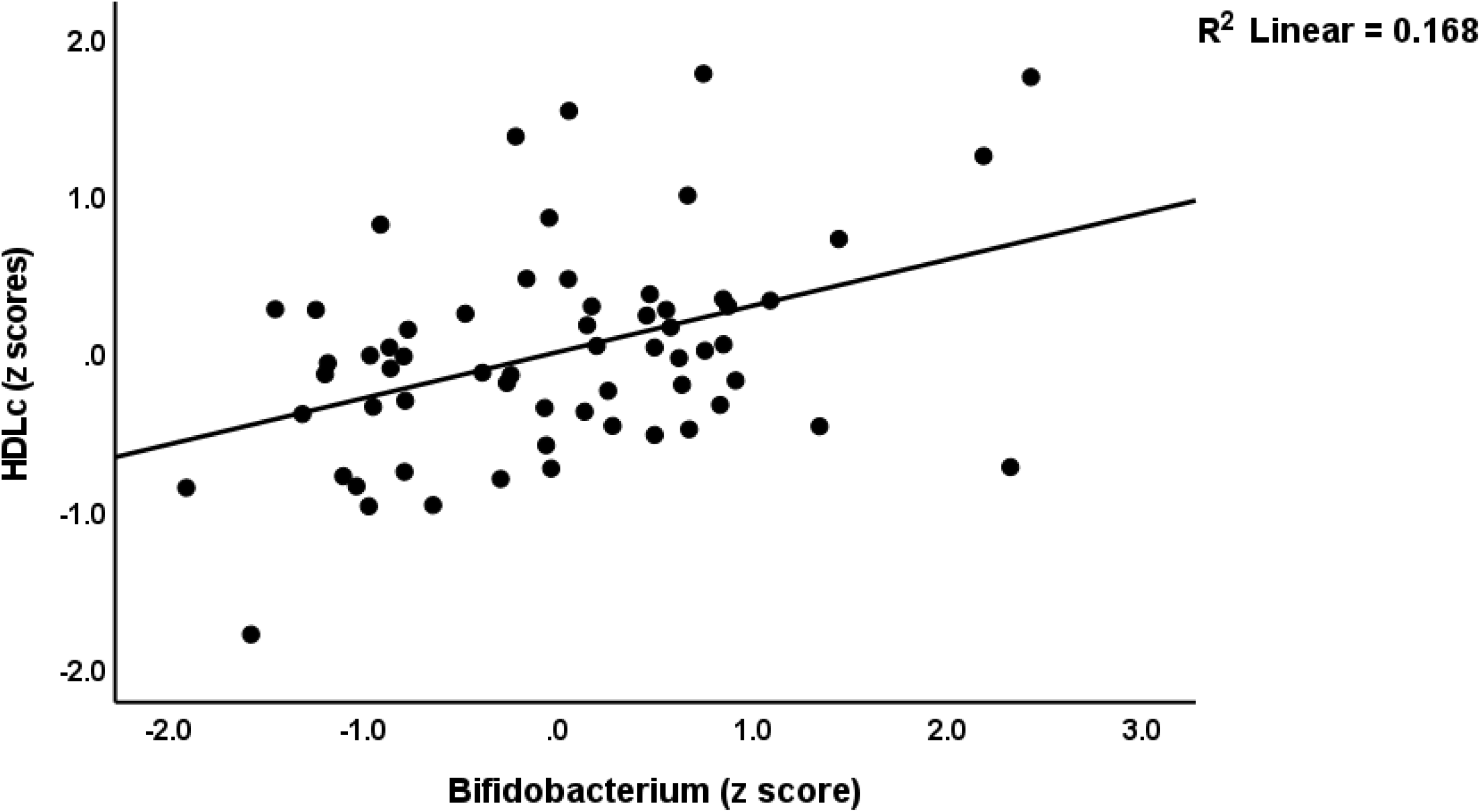
Partial regression of high-density lipoprotein cholesterol (HDLc) on the isometric log-ratio abundance of *Bifidobacterium*

Table 3 shows that this enterotype was not significantly different between controls and patients. Table 4 shows that enterotype 2 was significantly associated with PC_abuse but not with the number of episodes, LT_SB, ROI, PC_phenome, PC_Stroop, current_SB, and enterotype 1. Table 5, regression #6 shows that 45.8% of the variance in PC_phenome was explained by both enterotypes 1 and 2 and sex. **Figures 4 and 5** show the partial regressions of PC_phenome on enterotype 1 and 2, respectively. Enterotype 1 (p<0.001) and enterotype 2 (p=0.036) together explained 40.4% of the variance in the phenome (F=20.35, df=2/60, p<0.001).

**Figure 4.**
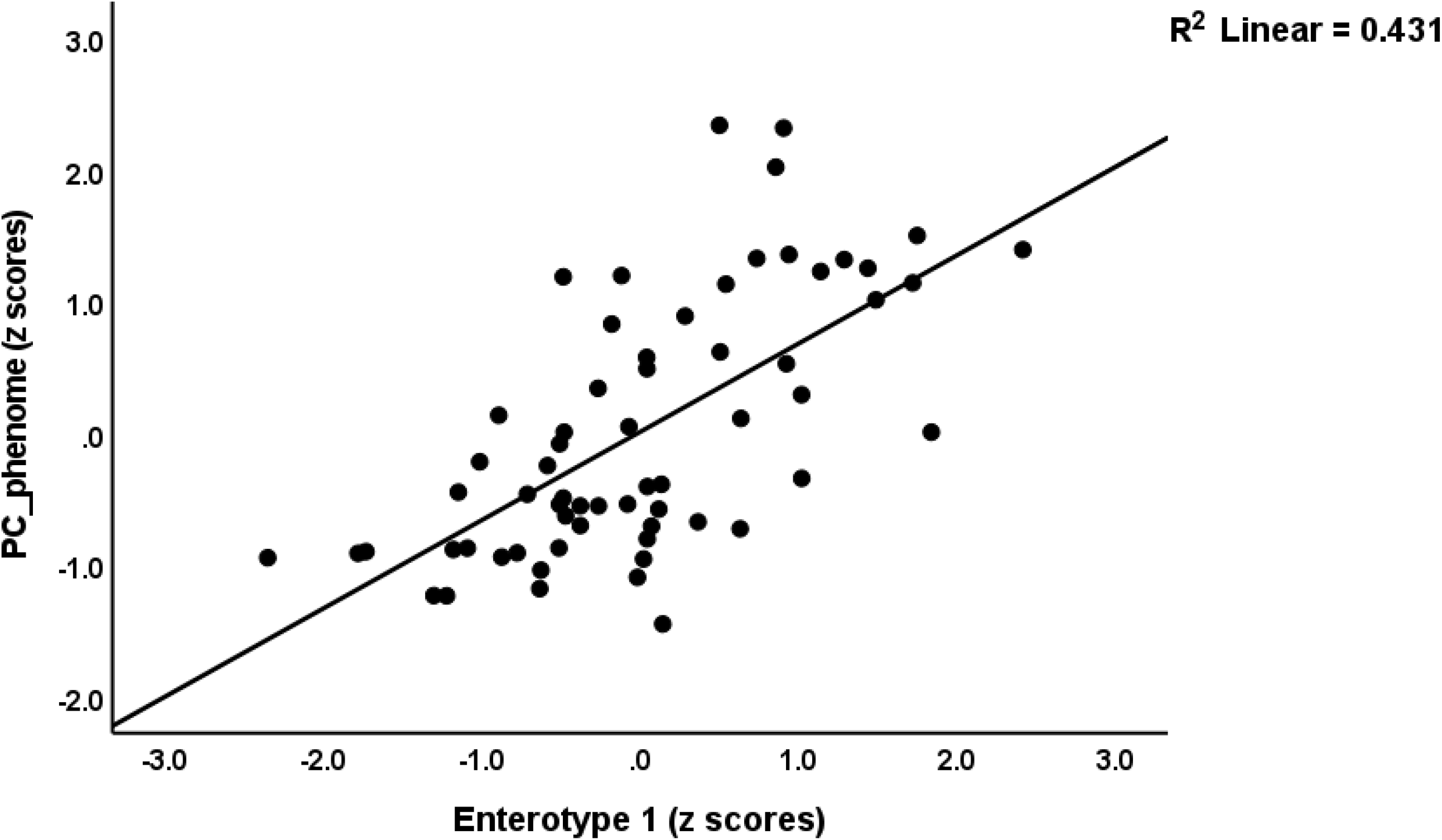
Partial regression of the phenome of depression (PC_phenome) on enterotype 1, a dysbiosis index of depression.

**Figure 5.**
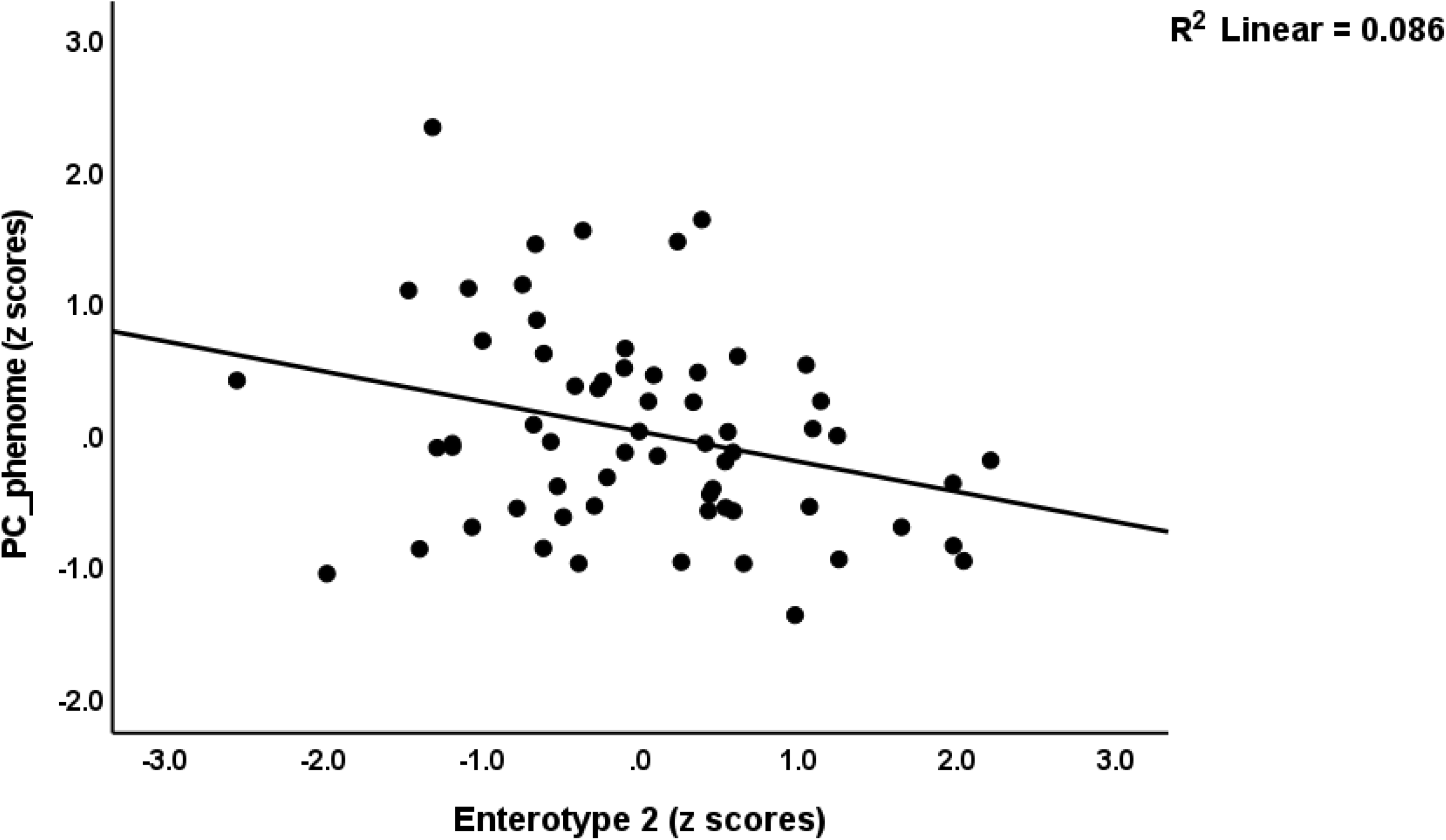
Partial regression of the phenome of depression (PC_phenome) on enterotype 2, a dysbiosis index of antioxidant-metabolic aberrations in depression.

Table 6 shows the outcome of linear modeling with overfit prevention with AIP as the dependent variable and the microbiome taxa as explanatory variables. Regressions #3 shows that *Acidaminococcus, Sutterella* and *Clostridium sensu stricto* were significantly and positively associated with increased AIP, whereas *Verrucomicrobia* and *Bifidobacterium* were inversely associated. AIP was significantly correlated with BMI (r=0.573, p<0.001) and enterotype 2 (r=-0.609, p<0.001). Both HDLc (r=-0.521, p<0.001) and enterotype 2 (r=-0.399, p=0.001, n=63) were significantly and inversely correlated with BMI. We were able to extract one PC from BMI, HDLc and enterotype 2 (KMO=0.627, Bartlett’s sphericity test χ2=58.691, df=3, p<0.001, VE=69.56%, all loadings >0.744).

### Results of PLS analysis

**Figure 6** shows the final PLS model after feature reduction (only the significant paths are shown). We entered two latent vectors, one reflecting the phenome (extracted from BDI, HAMD and Current_SB) and a second reflecting ROI (extracted from the number of episodes and LT_SB). All other variables were entered as simple indicators, whereby ROI, both enterotypes, HDLc and AIP, were allowed to mediate the effects of ACE on the phenome. The model quality criteria were adequate: SRMR = 0.040, and the extracted factors showed AVE values > 0.769 with Cronbach’s alpha > 0.732, composite reliability > 0.882, and rho_A > 0.732. PLS Blinfolding showed that the construct cross-validated redundancies were more than adequate, while PLS Predict showed sufficient model replicability. Complete PLS analysis, performed using 5,000 bootstraps, showed that 75.6% of the variance in the phenome was explained by enterotype 1, HDLc and ROI. The latter explained 28.5% of the variance in enterotype 1, whereas enterotype 2 explained 42.7% of the variance in HDLc. Consequently, enterotype 1 was a partial mediator of the effects of ROI on the phenome. Enterotype 2 showed a significant specific indirect effect on the phenome (p=0.038). PC_emabuse, PC_sexabuse and Comp_physneglect showed significant specific indirect effects on the phenome, which were mediated by ROI (p<0.001, p=0.004 and p=0.022, respectively) and the path from ROI to enterotype 1 (p=0.014, p=0.025 and p=0.042, respectively). PC_physabuse had no significant effect on the phenome (p=0.069) but affected the AIP (p=0.003).

**Figure 6.**
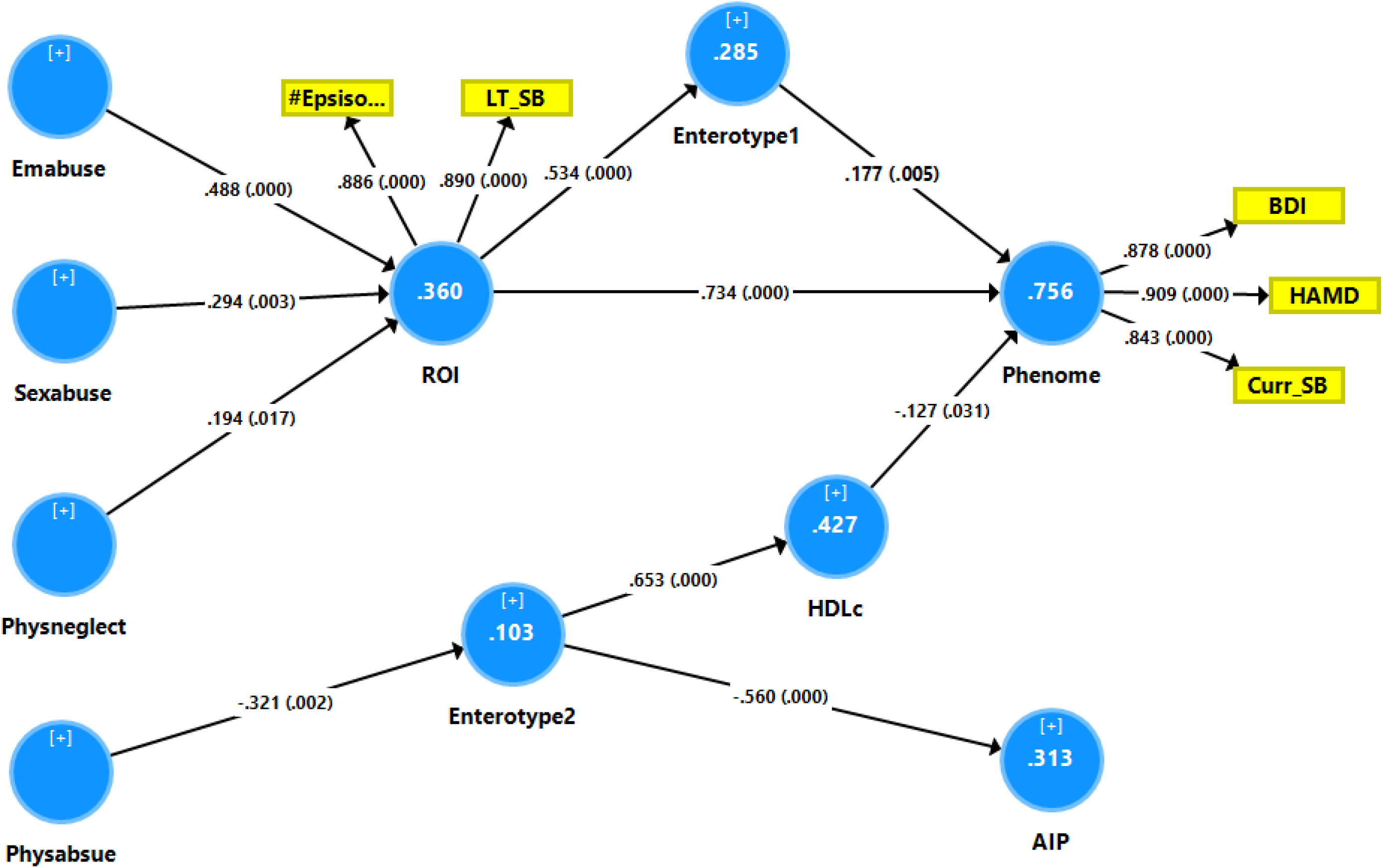
Results of Partial Least Squares (PLS) analysis. Phenome: first factor extracted from the BDI (Beck Depression Inventory) and HAMD (Hamilton Depression Rating Scale) scores and current suicidal behaviors (Curr_SB) scores; ROI: recurrence of illness; HDLc: high density lipoprotein cholesterol; enterotype 1 and 2: two dysbiosis indices, the first of the depressive phenome and the second of antioxidant-metabolic alterations in depression; emabuse: emotional abuse; sexabuse: sexual abuse; physneglect: physical neglect; physabuse: physical abuse. Shown are path coefficients with p-values of the inner model, and loadings with p-values of the outer model; figures in blue circles: explained variance.

## Discussion

### The enterotype of the phenome of depression

This study’s first key discovery is that the phenome of MDD is predicted by a composite of six microbiome taxa, designated enterotype 1, which collectively accounted for around 35- 36% of the variance. The major taxa contributing positively to this enterotype 1 are *Hungatella* (genus of anaerobic, Gram-positive bacterial; Sharma et al., 2019) and *Fusicatenibacter* (genus of anaerobic Gram-positive bacteria, Takada et al., 2013), whereas four other genera/species have inverse effects, namely *Butyricicoccus* (genus of anaerobic, Gram-positive bacteria, UniProt, 2023; Eeckhout et al., 2008), *Clostridium* (genus of anaerobic Gram-positive bacteria, Maczulak, 2011), *P. merdae (*species of anaerobic, Gram-negative bacteria, UniProt, 2023) and *D. piger (aerotolerant, Gram-negative bacterium*, Health Matters, 2022), Previously, using the same study group, we determined (via LEfSe analysis) the differences in relative abundance between MDD and controls. In accordance with the current analyses, *Hungatella hathewayi* (anaerobic, Gram-positive bacterium, Xia et al., 2020) was positively associated with MDD, while *D. piger* was inversely associated. Nevertheless, in our previous study, MDD was additionally associated with some other taxa. However, the phenome of depression assessed in our investigations (Maes, 2022; Maes et al., 2022a; Maes and Stoyanov, 2022) is a significantly more accurate measure of depression than the binary MDD diagnosis. The phenome evaluates the severity of the combination of several depressive features, and as a quantitative score, and provides more information than MDD, which is an incorrect model (see Introduction). Comparing the results of the present investigation conducted on Thai MDD patients with those of previous LefSe studies conducted in other cultures and nations reveals almost no agreement (Zhang et al. 2022; Zhao et al., 2022; Ling et al., 2022; Liu et al., 2022; Jiang et al., 2015; Zhu et al., 2021; Painold et al., 2018; Tsai et al., 2022). Notably, the LefSe study published by Liu et al. (2022) revealed a higher abundance of *Clostridium* in the control group, which is consistent with a lower abundance being related to the depressive phenome in the current study. In addition, there is limited consensus among all previously published investigations (see Introduction, Borkent et al., 2022). As described in our Introduction, this lack of consistency among studies may be explained by using the inaccurate diagnosis MDD (Maes, 2022) and by the knowledge that the composition of the microbiome is greatly influenced by nutrition (Singh et al., 2017). For instance, variations in the dietary inflammatory index elicit particular alterations in microbiome makeup (Costa et al., 2022). Therefore, it is likely that MDD enterotypes developed in one country will not coincide with those established in other nations. Deciphering whether the enterotype established here indicates compositional dysbiosis (see definition in the Introduction) is more important than just identifying a list of MDD-related taxa.

### Compositional dysbiosis and the phenome of depression

Four of the six microbiota taxa/species of enterotype 1 may promote salutogenesis; so, a reduction in their abundance may have negative implications. *Butyricicoccus* is a gut-mucosa-associated genus that appears to regulate the functioning of tight junctions (Devriese et al., 2017). Low levels of *Butyricicoccus* are associated with dysfunctions in tight junctions and inflammatory bowel disease (Devriese et al., 2017; Eeckhout et al., 2008). Intestinal butyrate improves the gut-immune defense barrier and mucosal inflammation and redox status, controls intestinal motility, energy consumption, neurogenesis, metabolic disorders such as atherogenicity, and insulin resistance (Canani et al., 2011). In addition, the *Clostridium* genus reduces inflammation, and numerous strains and species are key producers of CSFA, including butyrate, which inhibits ammonia absorption, supports Treg functions, and inhibits pathogen invasion (Guo et al., 2020). On the basis of these findings, Clostridium species have been recommended as potential probiotics for promoting gut health and ameliorating inflammatory bowel disease (Guo et al., 2020). The *Parabacteroides* genus and its various species produce SCFA, regulate the host’s metabolism, possess anti-inflammatory effects, and may strengthen the intestinal epithelium (Hiippala et al., 2020; Cui et al., 2022). *P. merdae* protects against cardiovascular diseases by, among other mechanisms, inhibiting the mTORC1 pathway and promoting the breakdown of branched-chain fatty acids (BCAA) (Qiao et al., 2022). As a consequence, *Parabacteroides* including *P. merdae* are presented as putative probiotics (Cui et al., 2022). *Bacteroides* and *Desulfovibrio* genera, including *D. piger*, are sulfate-reducing bacteria (SRB) and are sulfidogenic, namely they convert sulfur-containing substrates (e.g., cysteine) to hydrogen sulfide (Nguyen et al., 2020; Loubinoux et al., 2003). Hydrogen sulfide at low concentrations is protective and maintains mucus layer integrity, has anti-inflammatory properties, aids in the resolution of tissue damage, prevents the adhesion of microbiota biofilms to the epithelium, and inhibits invasive pathobionts (Blanchier et al., 2019; Buret et al., 2022; Dordevic et al., 2020). Additionally, hydrogen disulfide produced from the gut has cardioprotective properties, promotes vasodilation and reduces the heart rate (Tomasova et al., 2016).

Two microbiota genera in enterotype 1 may have pathophysiological effects, in contrast. First, the *Hungatella* genera and *H. hathewayi* (as identified in our LefSe study, Maes et al., 2022c) are potential pathogens related to cardiovascular illness, Chrohn’s disease, and colorectal cancer (Kaur et al., 2014; Human Gut Microbiome Atlas, 2023). In addition, *Hungatella* is one of the genera that creates trimethylamine (TMA), a uremic toxin and precursor of trimethyl-N-oxide (TMAO), from choline, carnitine, and betaine present in meat, eggs, and shellfish (Genoni et al., 2020; Macpherson et al., 2020). After being delivered to the liver, TMA is oxidized into TMAO, which may trigger systemic inflammation via increased production of cytokines such as interleukin (IL)-12 and tumor necrosis factor (TNF)-α, and is accompanied by increased gut permeability (as demonstrated by elevated plasma LPS) (Macpherson et al., 2020). In some circumstances, the increase in TMA-producing genera (such as *Hungatella*) is followed by a decrease in *Bifidobactrium* (Macpherson et al., 2020).

*Fusicatenibacter* is prevalent in insomnia sufferers, despite its anti-inflammatory properties and decreased prevalence in inflammatory illnesses (Zhou et al., 2022; Zanelli et al., 2005; Lee 2019). *Fusicatenibacter* is nonetheless a glucose fermenter that generates acetic acid, succinic acid, formic acid, and lactic acid (Midas Field Guide, 2023; Takada et al., 2013).

Lactate has multiple impacts on the immune system and inflammatory response, including actions on the G-protein coupled receptor and NF-κB (Manosalva et al., 2022). Due to decreased tissue oxygenation and deficiencies in mitochondrial respiration, elevated lactate levels are observed in depressed phenotypes and chronic fatigue (Morris and Maes, 2013; Machado-Vieira et al., 2017). Therefore, increased gut-derived lactic acid levels could exacerbate elevated lactate in MDD and hence exacerbate depressive symptoms (Chen et al., 2022). Formic acid may impede mitochondrial cytochrome oxidase and ATP synthesis, activate oxidative stress responses, T helper-17 responses, and the aryl hydrocarbon pathway, and so exacerbate systemic metabolic acidosis (Liesivuori and Savolainen, 1991; Ternes et al., 2022). In addition, because formic acid has direct decontaminating effects on Gram-negative bacteria, it may lead to microbiota imbalances. Succinate signaling is essential for metabolic activities, the Krebs cycle, and cell-to-cell communication, as well as chemotaxis and T-cell activation, while its receptor (SUCNR1) synergizes with the TLR to promote the production of pro-inflammatory cytokines such as IL-1β and TNF-α (Tretter et al., 2016; Mills and O’Neill, 2014). Generally speaking, acetic acid is a mildly toxic chemical that promotes mixed lymphocyte and natural killer cell reactivity (Ishizaka et al., 1993) and directs immune cells toward an immunological defense response (Balmer et al., 2020).

As a consequence, the enterotype 1 identified in this study may imply compositional dysbiosis with diminished salutogenesis (decreased butyric acid and hydrogen disulfide synthesis, diminished gut-immune protection against inflammation and oxidative stress) and increased pathogenesis (increased formic, acetic, lactic acid and TMAO production, breakdown of the gut barrier, LPS translocation and inflammation). Consequently, we have developed an enterotype dysbiosis index of the phenome of depression.

### Enterotype 1, ACEs, ROI, suicidal behaviours and neurocognition

The second significant finding of our study is that enterotype 1 is affected by childhood abuse and is so closely linked with ROI and lifetime suicidal behaviors that a latent vector could be extracted, reflecting a ROI-enterotype pathway phenotype. In addition, we were able to extract one latent vector from abuse, ROI, the phenome, and the enterotype 1 dysbiosis index, demonstrating that compositional dysbiosis is a key component of depression’s lifespan trajectory (from ACEs to ROI to the phenome).

According to a preliminary study, ACEs may elicit changes in gut microbiome composition during pregnancy, contributing to systemic inflammatory and hypothalamic-pituitary-adrenal-axis responses (Hantsoo et al., 2019). Recently, we discovered that ACE is connected with sensitized immunological and growth factor networks, nitro-oxidative stress, and antioxidant pathways (Maes et al., 2019; 2022b; Moraes et al., 2018). Consequently, it was postulated that the microimmuneoxysome is a potential therapeutic target for deprogramming the negative effects of ACEs (Dietert and Dietert, 2022). To the best of our knowledge, no research has linked changes in the microbiome to recurrent suicidal ideation or behaviours. However, earlier studies demonstrated that leaky gut indicators were connected with suicidal behaviours (Ohlsson et al., 2019). In addition, we determined that, apart from enterotype 1, suicidal behaviours were also connected with the abundance of *Proteobacteria*. The latter phylum contains numerous pathogens that can induce intestinal (e.g., inflammatory gut disease) and metabolic diseases, in addition to lung diseases (Rizzati et al., 2017). Additionally, the prevalence of *Proteobacteria* is linked to inflammatory reactions, elevated IgA levels, and TMA production (Li et al., 2021).

Deficits in the Stroop test (showing dysfunctions in processing speed, cognitive flexibility, selective attention, and executive functioning) were related to an increase in the abundance of *Dialister* and a decrease in *Intestinimonas* abundance. The latter is a butyrate-producing genus that may protect against type 2 diabetes (Bui et al., 2020; NIH Clininal Trials, 2023). *Dialister* is a possible gut dysbiosis marker in inflammatory bowel disease, ulcerative colitis, and spondylarthritis (Tito et al., 2017; Nwosu, 2011).

### Enterotype 2, metabolism, and the phenome of depression

The third significant discovery of this study is that we were able to create a second enterotype that reflects changes in HDLc and, consequently, AIP and BMI. *Bifidobacterium, P. merdae*, and *Romboutsia* were positively correlated with HDLc, whereas *Proteobacteria* and *Clostridium sensu stricto* were negatively correlated. *Bifidobacterium* is a protective genus that supports the gut barrier and gut homeostasis protects against the multiplication of pathogens, and produces SCFAs, vitamins, and polyphenols (Alessandri et al., 2021). Moreover, *Bifidobacterium* has antiobesity and cholesterol-reducing actions (An et al., 2011) and is associated with leanness (Xu et al., 2022). As mentioned previously, *P. merdae* has numerous health-supporting properties, whilst this species has been advocated for weight, body fat, and triglyceride reduction (TWI609959B, 2016). The *Romboutsia* genus produces SCFAs and many metabolic end products based on carbohydrate utilization and amino-acid fermentation (Gerritsen 2015). *Proteobacteria* are the most consistently reported microbiota related to obesity in the aforementioned systematic research (Xu et al., 2022). *Clostridium sensu stricto* is a putative opportunistic pathogen that can lead to decreased SCFA levels and intestinal inflammation (Hu et al., 2021). In swine, correlation heat map analysis demonstrated that *Clostridium sensu stricto* is strongly connected with total cholesterol and the pathogenesis of heat-stress-associated inflammatory bowel disease (Hu et al., 2021).

*Bifidobacterium* (in a negative direction) and *Clostridium sensu stricto* (in a positive direction) were also predictors of AIP, which was also associated with decreased abundance of *Verrucomicrobia* and an increased abundance of *Acidaminococcus* and *Sutterella*. *Verrucomicrobia* is a phylum that promotes gut health, gut barrier function, and insulin sensitivity and inhibits inflammatory responses (Fujio-Vejar et al., 2017). Obese people have a lower incidence of *Verrucomicrobia* (Zhang et al., 2009). The presence of *Sutterella* is associated with inflammatory bowel disease (Eid et al., 2017; Williams et al., 2012) and is a potential initiator of T2DM (Gradisteanu Pircalabioru et al., 2022). The relative abundance of *Acidaminococcu*s is positively associated with obesity in Italian adults (Palmas et al., 2021).

Importantly, we discovered that both enterotypes predicted the depression phenome and that the latter was inversely associated with HDLc. As a result, we have developed an enterotype dysbiosis index that reflects decreased HDLc, which is a strong predictor of the antioxidant defenses against lipid peroxidation (Maes et al., 2021), increased atherogenicity and elevated BMI, and consequently obesity (Morelli et al., 2021). As with enterotype 1, the second compositional dysbiosis index was associated with childhood abuse, but the effect size was much smaller. We have previously demonstrated that ACE, and particularly sexual abuse, impact antioxidant defences (Maes et al., 2021). In this regard, our PLS pathway analysis revealed that diverse ACEs influence the phenome of depression and current suicidal behaviors, and that these effects are mediated by the two gut dysbiosis indices. Moreover, HDLc was inversely associated with current, but not lifetime, suicidal behaviors. Previously, we have shown that lowered HDLc is associated with suicidal attempts in depressed patients (Maes et al., 1997).

## Limitations

This study would have been more intriguing if oxidative stress biomarkers had been measured in addition to the immune and growth factor networks. It could be argued that the study’s sample size and statistical power are low. Nevertheless, an a priori calculation of the sample size revealed that a sample size of n=65 is required to achieve a power of 0.80. Moreover, the regression of the phenomes of the six microbiota of enterotypes 1 and 2 revealed that, given the study sample, alpha=0.05, and 5-6 predictors, the obtained power was 0.995 and 0.992, respectively. A previous COVID-19 infection is yet another possible intervening factor through the onset of Long-COVID. However, we excluded all participants with moderate and severe COVID-19, as these are the types predisposing to Long-COVID affective disorders (Al-Hadrawi et al., 2022). In addition, there were no significant effects of previous (at least 6 months before enrollment) mild COVID-19 on the microbiome or clinical data. Both enterotypes 1 and 2 ought to be cross-validated in a new Thai study population. Future research should construct region- and culture-specific dysbiosis indices of ROI, the phenome, suicidal behaviors, cognitive deficits, and metabolic abnormalities of depression.

## Conclusions

Six microbiome taxa, including positive associations with *Hungatella* and *Fusicatenibacter* and negative associations with *Butyricicoccus, Clostridium, P.* merdae, and *D. piger*, accounted for 36% of the variance in the depression phenome. Based on these data we constructed a composite score, namely enterotype 1, indicative of compositional dysbiosis. Enterotype 1 is strongly predicted by ACE and ROI, and is associated with suicidal behaviours. We constructed another enterotype 2 that reflects a decrease in HDLc and an increase in AIP based on *Bifidobacterium, P. merdae, and Romboutsia* (positively associated with HDLc), and *Proteobacteria* and *Clostridium sensu stricto* (inversely associated with HDLc). Together, enterotypes 1 and 2 accounted for 40.4% of the variance in the depression phenome, and enterotype 1 in combination with HDLc accounted for 39.9% of the variance in current suicidal behaviours. In conclusion, both enterotypes are potential new drug targets for the treatment of severe depression and suicidal behaviours, as well as the possible prevention of future episodes. Moreover, the “microimmuneoxysome” is a new drug target for “desensitizing” the ROI and “deprogramming” the effects of ACE, leading to increased ROI and severity of the phenome and suicidal behaviors. Future research should trial the therapeutical effects of butyrate supplements, zinc and glutamine (Maes et al., 2007) as well as probiotic preparates with *Clostridium* species to improve the features of depression, including ROI, suicidal behaviors and the phenome in association with enterotype and leaky gut assessments. In addition, the development of new drugs targeting leaky gut would be more than welcome.

## Data Availability

The dataset generated during and/or analyzed during the current study will be available from MM upon reasonable request and once the authors have fully exploited the dataset.

## Author Declarations

### Author’s Contributions

All authors contributed to the paper. MM designed the study. Patients were recruited by AV, KJ and CT. Microbiome assays were performed by PV, PC and SP. Statistical analyses were performed by M.M. Abundance data were transformed by RP and AS. All authors revised and approved the final draft.

## Compliance with Ethical Standards

### Research involving Human Participants and/or Animals

This study was approved by the Institutional Review Board (IRB) of Chulalongkorn University, Bangkok, Thailand (IRB no. 62/073), which complies with the International Guideline for Human Research Protection as required by the Declaration of Helsinki.

### Informed consent

Before taking part in the study, all participants and/or their caregivers provided written informed consent.

